# Estimating the impact of COVID-19 control measures using a Bayesian model of physical distancing

**DOI:** 10.1101/2020.04.17.20070086

**Authors:** Sean C. Anderson, Andrew M. Edwards, Madi Yerlanov, Nicola Mulberry, Jessica E. Stockdale, Sarafa A. Iyaniwura, Rebeca C. Falcao, Michael C. Otterstatter, Michael A. Irvine, Naveed Z. Janjua, Daniel Coombs, Caroline Colijn

## Abstract

Extensive physical distancing measures are currently the primary intervention against coronavirus disease 2019 (COVID-19) worldwide. It is therefore urgent to estimate the impact such measures are having. We introduce a Bayesian epidemiological model in which a proportion of individuals are willing and able to participate in distancing measures, with the timing of these measures informed by survey data on attitudes to distancing and COVID-19. We fit our model to reported COVID-19 cases in British Columbia, Canada, using an observation model that accounts for both underestimation and the delay between symptom onset and reporting. We estimate the impact that physical distancing (also known as social distancing) has had on the contact rate and examine the projected impact of relaxing distancing measures. We find that distancing has had a strong impact, consistent with declines in reported cases and in hospitalization and intensive care unit numbers. We estimate that approximately 0.78 (0.66–0.89 90% CI) of contacts have been removed for individuals in British Columbia practising physical distancing and that this fraction is above the threshold of 0.45 at which prevalence is expected to grow. However, relaxing distancing measures beyond this threshold re-starts rapid exponential growth. Because the extent of underestimation is unknown, the data are consistent with a wide range in the prevalence of COVID-19 in the population; changes to testing criteria over time introduce additional uncertainty. Our projections indicate that intermittent distancing measures—if sufficiently strong and robustly followed— could control COVID-19 transmission, but that if distancing measures are relaxed too much, the epidemic curve would grow to high prevalence.

## 1 Introduction

Coronavirus disease 2019 (COVID-19), caused by the severe acute respiratory syndrome coronavirus 2 (SARS-CoV-2), has now spread worldwide and resulted in over 1.5 million diagnosed cases and 100,000 deaths globally [1]. Estimates of COVID-19 case fatality rates from Hubei, China, the rest of China, and other countries range from 0.3% to above 5% in different populations at various times [2, 3], with an estimate of 1.4% [4] currently favoured in some analyses [5]. Age [6] and comorbidities (hypertension, cardiovascular and respiratory disease [7]) are strong risk factors for severe illness, hospitalization, and death. Furthermore, COVID-19 poses severe challenges for health care, with risks that requirements will exceed hospital bed, critical care and ICU capacities even in well-resourced health care systems. In the current absence of a vaccine or effective therapeutic options, widespread physical distancing measures (also known as social distancing measures) and handwashing are the main interventions currently available to reduce transmission. Countries have used a variety of such physical or social distancing measures including cancelling mass gatherings, closing restaurants, work-from-home orders, and “lockdowns” of varying strictness.

In British Columbia (BC), Canada, the first case of infection was detected on January 26, 2020 with sporadic cases related to travel until March 8, followed by a sustained increase in cases. A number of measures were implemented over the following weeks to reduce transmission (Figure 1). However, the direct impact of these measures on transmission is not known. Distancing measures have high economic, health, and social impacts. Thus, there is an urgent need to understand what level of contact rate and physical distancing measures are optimal to reduce transmission. Once initial transmission has been brought under control, as in China and Korea [8, 9], there remains the question of what relaxation in social measures could keep transmission under control.

**Figure 1:**
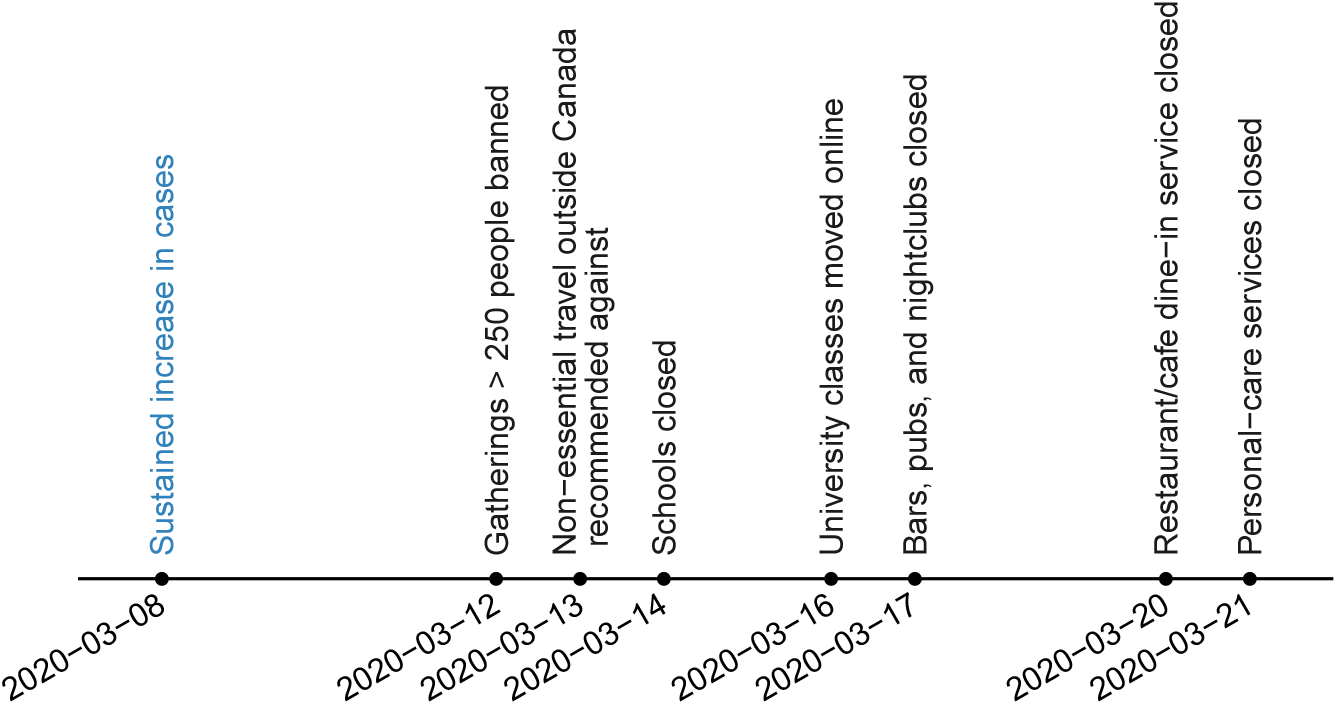
Key physical distancing measures implemented in British Columbia, Canada in response to COVID-19. Schools closed for an annual two-week break on March 14, then were declared indefinitely closed on March 17.

There have been a number of models simulating the impact of broad physical distancing measures [8, 10, 11, 12]. Direct estimates of the strength and impact of distancing measures have focused on the effective reproduction number over time, using approaches based on reported deaths [13] or confirmed cases [14]. These estimates are influenced by the assumed serial interval distribution and the infection fatality and delay between symptom onset and death. In comparisons among EU countries, estimates assume that physical-distancing measures impact each location equally and that all deaths are reported [13]. Inferences from reported deaths also impacted by the assumption that deaths are independent, whereas in British Columbia, more than half are associated with clusters in long term care facilities. Estimates of the effective reproduction number based on reported cases have been adjusted for the delay between symptom onset and reporting [14, 15], but cannot accommodate underestimation or asymptomatic or weakly symptomatic individuals. Furthermore, the effective reproduction number is a broad summary of the overall growth of the epidemic, and is not a direct estimate of the impact of physical distancing on the contact patterns relevant to transmission.

Here, we introduce an epidemiological model of physical distancing and assess the degree to which contact rates have changed—for the population that is participating in physical distancing— due to recent policy measures in British Columbia. The model structure is informed by a survey of attitudes and responses to messaging about COVID-19 and physical distancing. We use an observation model to reflect the relationship between case counts and incident infections and to account for underestimation of the time between symptom onset and case reporting. We then use a Bayesian approach to estimate the reduction in contact among those participating in physical distancing measures, using reported case counts for the period March 1 to April 11, 2020. We explore to what extent distancing measures could be relaxed while keeping case numbers under control.

## 2 Methods

### 2.1 Data

We fit the physical distancing model to case count data from British Columbia from March 1, 2020 (when a total of eight cases had been detected in the province) to April 11, 2020 at which time 1445 cases had been confirmed. These data are available in press releases from the BC Centre for Disease Control (BCCDC) [16], from the public data dashboard [17], and from the code repository associated with this paper. Testing procedures were adapted over the course of the outbreak. In particular, lab testing criteria were changed on March 16 to focus on hospitalized patients, healthcare workers, long term care facility residents/staff, and those people part of an existing cluster and experiencing respiratory symptoms. This led to instability in case counts in the surrounding days with some large jumps in the number of identified cases, which we correct for in the model. There was also considerable variability in the daily testing rate. During March, the daily number of completed tests ranged from approximately 100 to 3500, and did not strictly increase over time.

For some of the confirmed cases, estimates of the date of symptom onset are available (Figure 2). We used the delays between symptom onset and cases being reported to parameterize the physical distancing model. In this case-specific data set there are only seven cases reported before February 29, and a decline in reported cases after April 2 (Figure 2). Therefore we used only the 535 cases in the case-specific data that were reported between these dates to parameterize the delay part of the model.

**Figure 2:**
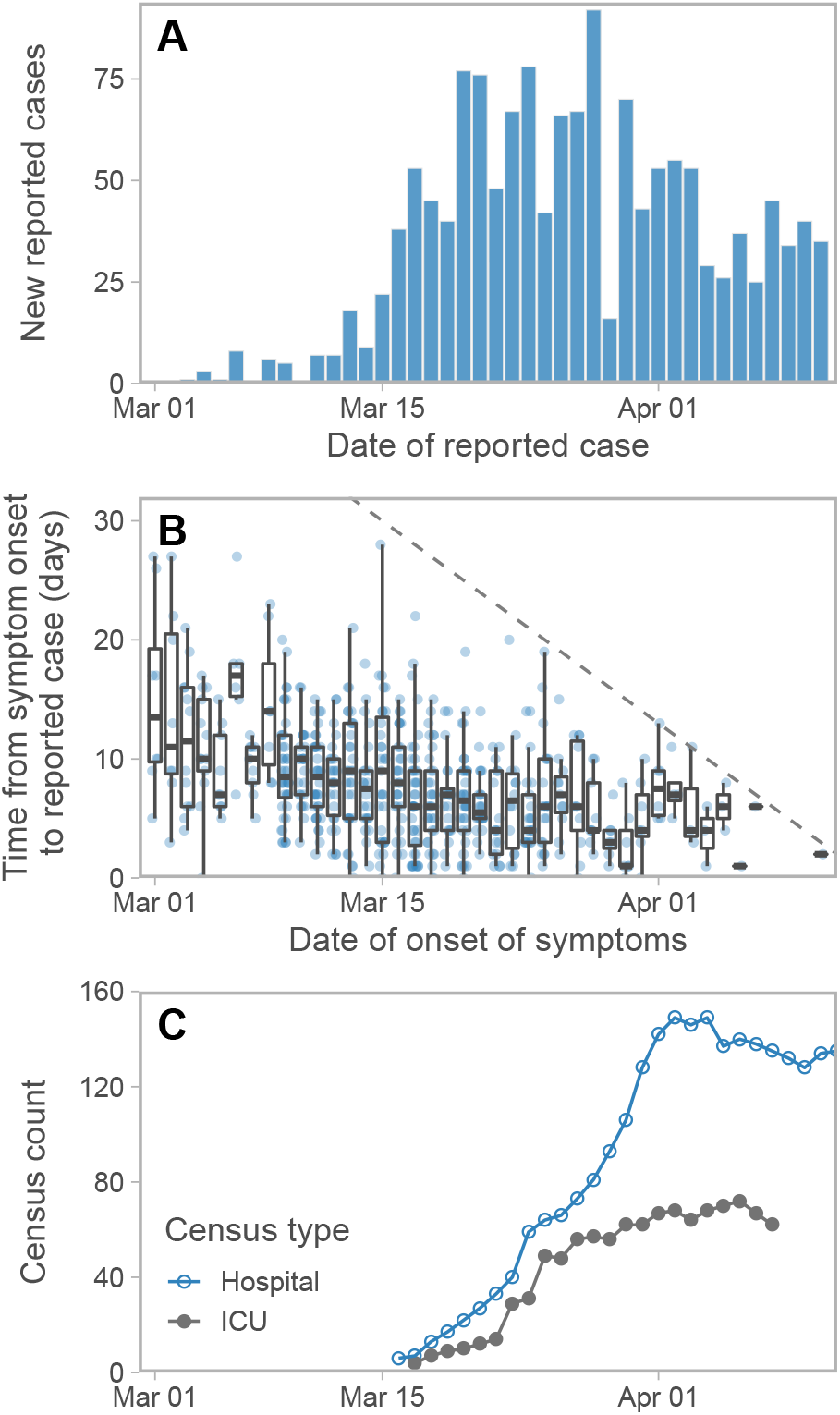
(A) New reported cases per day and (B) time from symptom onset to reporting from the case-specific data as of April 11, 2020. The dashed line in panel B represents the 1:1 line. Cases above this line, by definition, have not been reported yet. (C) Hospitalization and ICU (Intensive Care Unit) census counts. All data are from the BC Centre for Disease Control.

The Angus Reid Institute conducted a survey to examine how physical distancing measures have changed behaviour (March 20–23, 2020; n = 1664; [18]). The survey questions included whether respondents believed the COVID-19 epidemic was “overblown”. Responses about behavioural practices around hand washing, keeping extra distance, and other distancing behaviours were stratified according to this belief.

### 2.2 Epidemiological model

We developed an epidemiological model to describe the numbers of individuals who are: susceptible (*5*); exposed to the virus (*E*_1_); exposed, pre-symptomatic, and infectious (*E*_2_); symptomatic and infectious (*I*); quarantined (*Q*); and recovered or deceased (*R*) over time (Figure 3). Recovered individuals are assumed to be immune. The model includes analogous variables for individuals practising physical distancing, represented by subscript d, i.e. *5*_d_, *E*_1d_, *E*_2d_, *I*_d_, *Q*_d_, and *R*_d_. Physical distancing reduces the frequency of contact between individuals (and hence between compartments; Figure 3).

**Figure 3:**
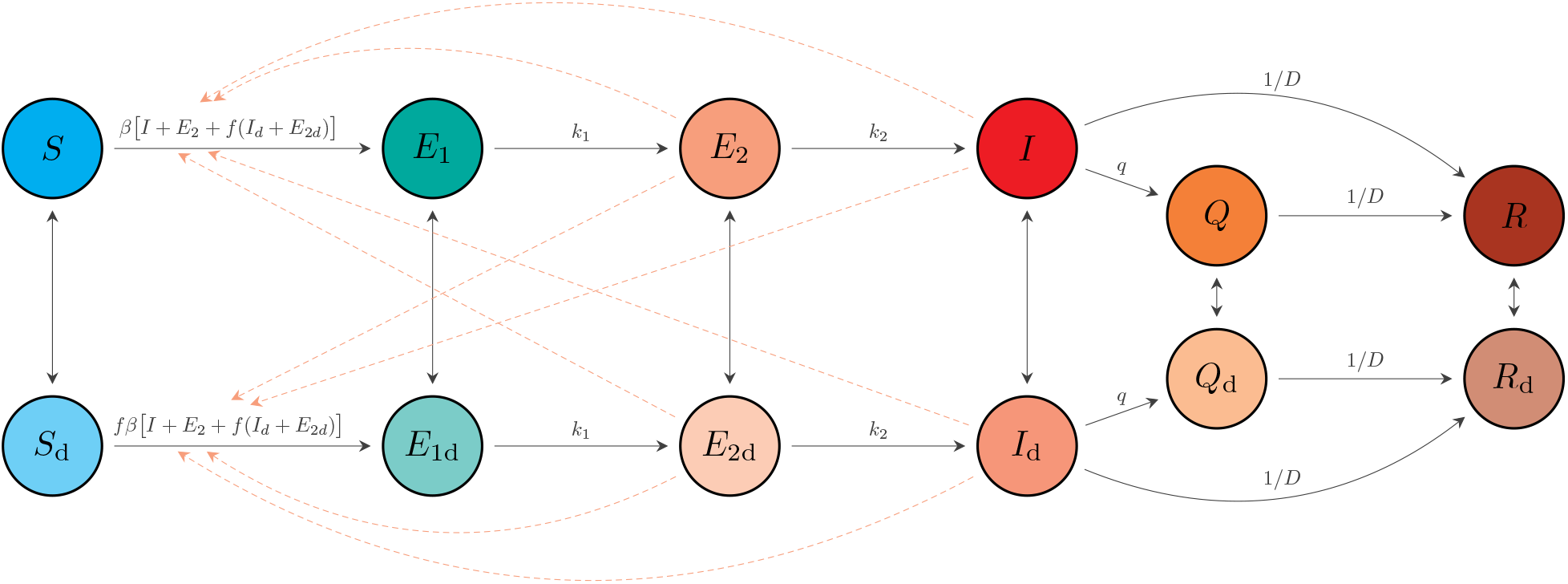
Schematic of model described by Equations (1) and (2). An individual in some compartment *X* can begin distancing and move to the corresponding compartment *X*_d_ at rate *u*_d_. The reverse transition occurs at rate *u*_*r*_. The model quickly settles on a fraction *e* = *u*_*d*_/(*u*_*d*_ +*u*_*r*_) participating in distancing, and dynamics depend on this fraction, rather than on the rates *u*_*d*_ and *u*_*r*_.

The non-physical-distancing differential equations are:

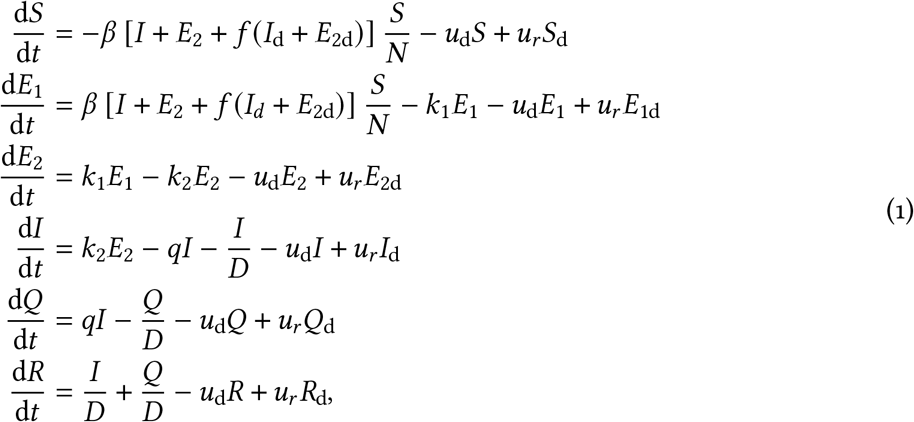

where *β* is the transmission parameter, *D* is the mean duration of the infectious period, *f* represents physical distancing, *u*_d_ and *u*_*r*_ are the rates of movement to and from the physical distancing compartments, *k*_1_ is the rate of movement from the *E*_1_ to *E*_2_ compartment, *k*_2_ is the rate of movement from the *E*_2_ to *I* compartment, and *q* is the quarantine rate for movement from the *I* to *Q* compartment. The physical distancing compartments contribute a reduced amount (a fraction *f*) to the force of infection, with *f* = 1 representing no physical distancing. We make *f* time-dependent (see below) to represent changes in physical distancing. We report the estimated impact of distancing in terms of the fraction of contacts that are reduced (i.e., 1− *f*), so that “strong distancing” is a high fraction (or percent), and “weak distancing” is a low fraction (or percent). In the model without interventions (neither distancing nor quarantine), the basic reproductive number *R*_0b_ is *β* (*D*+ 1/*k*_2_), namely the transmission rate times the mean duration in the infectious compartment.

There are six analogous equations for the physical distancing compartments (denoted by subscript d). The physically distancing compartment *5*_*d*_ has a contact rate with infectious individuals that is a fraction *f* of that for the *5* compartment. This factor appears twice: distancing individuals contribute less to the force of infection and also are less likely to encounter others than non-distancing individuals, giving

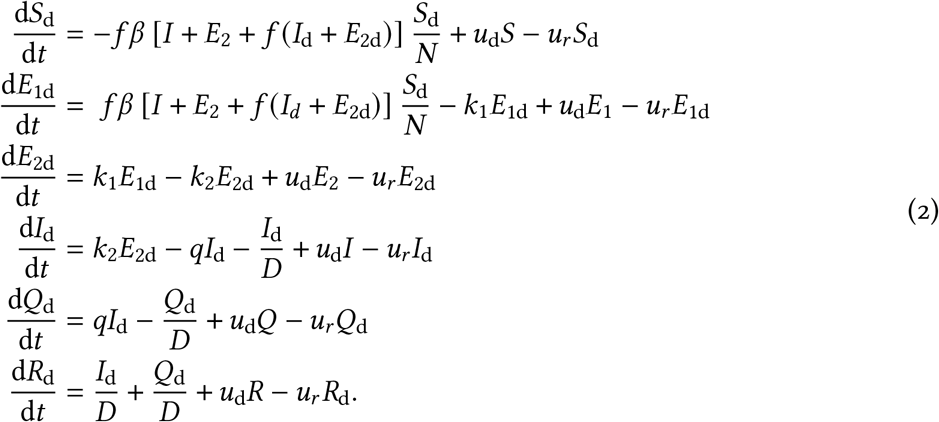

We model the change in physical distancing that occurred in British Columbia in March 2020 by a simple linear function whereby physical distancing increases (*f*(*t*) decreases) over one week between 15th March (*t* = *t*_1_) and 22nd March (*t* = *t*_2_):

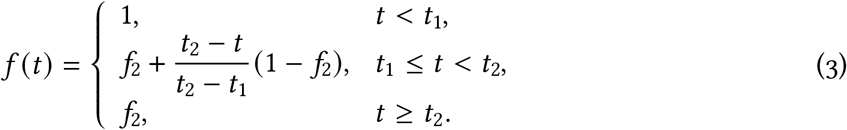

We fix *f* (*t*) = *f*_2_ until the final day of observed data and change *f* (*t*)for future days to represent different scenarios regarding relaxation of physical distancing.

The number of reported cases each day is *C*_*r*_, where *r* represents discrete days (whereas *t* is continuous time in the model). We also have data on each individual case, some of which have an estimated date of the onset of symptoms. The number of people per day who become symptomatic is the number per day moving from the *E*_2_ and *E*_2*d*_ compartments to the *I* and *I*_*d*_ compartments.

Due to the delay between symptom onset and reporting, the model’s predicted number of reported cases, *µ*_*r*_, on integer day *r* is comprised of contributions from previous days weighted by a delay:

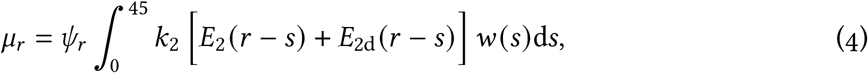

where *w* (*s*) is the density function for delay *s* and 45 is the maximum delay that needs to be considered (see Supplement). Also, *ψ*_*r*_ is the proportion of anticipated cases on day *r* that are tested and reported; this varies over time due to changes in the testing protocols, lab capacity and other factors. In our data, the number of tests performed each day jumped dramatically on March 14; we model this with a sharp increase in *ψ*_*r*_ on that date (0.1 to 0.3) and attempt to fit a random walk to this function as a supplementary analysis.

We use a Weibull distribution for the delay function *w* (*s*), and fit the shape and scale parameters using the case-specific data of reported cases and time of symptom onset (Figure 2B). In the Supplement we derive a novel multinomial likelihood function (based on [19] and [20]) to handle right-truncation. The resulting maximum likelihood estimate (with 95% univariate confidence interval) of the shape is 1.73 (1.60–1.86) and of the scale is 9.85 (9.30–10.46). Using the maximum likelihood estimates, the mean delay from symptom onset to reporting is 8.78 days, one day longer than the data’s mean (7.79 days) due to accounting for right-truncation (Figure S2).

### 2.3 Fixed parameter and variable values

The nature of the epidemiological model requires us to assume a number of parameter and variable values (Table 1). We test the robustness of our results to the key fixed values as supplementary analyses. Specifying initial conditions for the epidemiological model involves setting a total number of infected people *I*_0_ at an initial point in time (February 1, 2020), which are divided among the exposed and infected compartments. We define *e* = *u*_d_ / (*u*_d_+ *u*_*r*_) to be the fraction practising physical distancing (*e* = 0.83 in the base model). To further avoid sharp initial transient behaviour we also initially distribute all individuals among the distancing and non-distancing components, to give reasonable conditions for March 1 (Table 2). The model begins on February 1 with 8 cases, reflecting a 10–30% chance of detection [5] and very low reported cases at that time. We do not model introductions during February, and instead we compensate for this with an elevated initial number of infectious cases.

**Table 1:** Parameters and sources. The duration of the infectious period is shorter than the duration of severe illness, accounting for self-isolation and less severe illnesses. The quarantine parameter *q* reflects approximately 1/5 of severe cases either ceasing to transmit due to hospitalization or completely self-isolating. The model depends on the combination *u*_*r*_/(*u*_*r*_+ *u*_*d*_), the fraction engaged in physical distancing, estimated from the survey data cited above. The testing patterns have changed over time, with laboratories increasing the numbers of tests on approximately March 14 (motivating our change in *ψ*_*r*_).

| Symbol | Definition | Default value | Justification |
| --- | --- | --- | --- |
| $N$ | Population of British Columbia | 5,100,000 | [21] |
| $D$ | Mean duration of the infectious period | 5 days | [22, 23] |
| $k_1$ | (time to infectiousness)$^{-1}$ ($E_1$ to $E_2$) | 0.2 days$^{-1}$ | [24, 25, 26] |
| $k_2$ | (time period of pre-symptomatic transmissibility)$^{-1}$ ($E_2$ to $I$) | 1 days$^{-1}$ | [25, 26] |
| $q$ | Quarantine rate | 0.05 | [27] |
| $u_d$ | Rate of people moving to physical distancing | 0.1 | [18] |
| $u_r$ | Rate of people returning from physical distancing | 0.02 | [18] |
| $\psi_r$ | Proportion of anticipated cases on day $r$ that are tested and reported | 0.1 | Before March 14 |
|  |  | 0.3 | On and after March 14 |

**Table 2:**
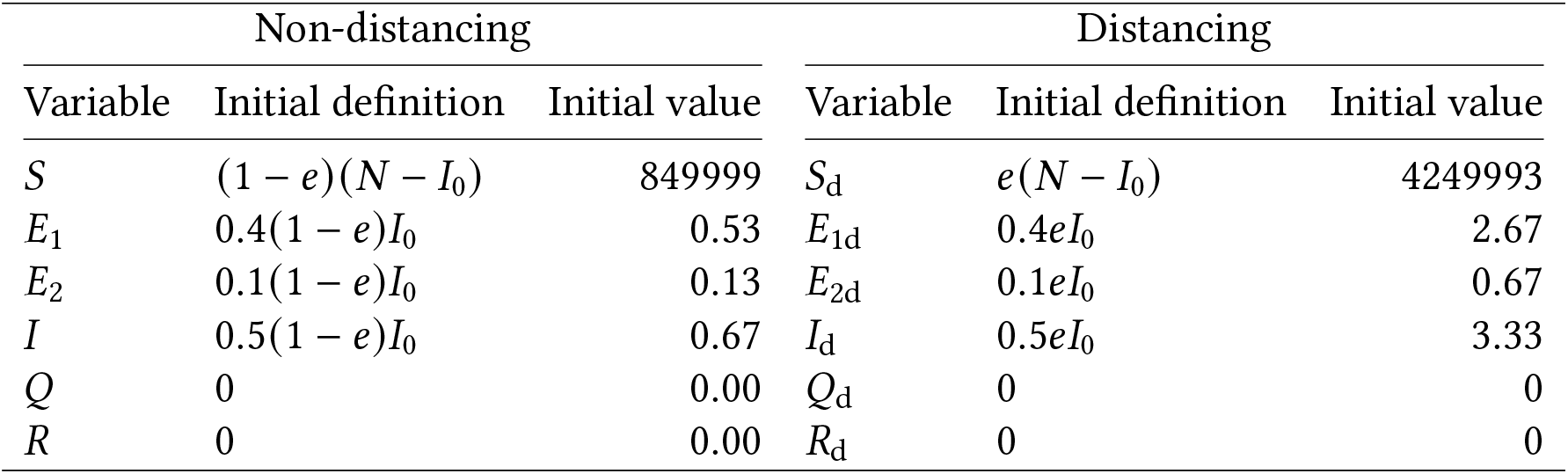
Initial values of variables for February 1, 2020.

| Non-distancing |  |  | Distancing |  |  |
| --- | --- | --- | --- | --- | --- |
| Variable | Initial definition | Initial value | Variable | Initial definition | Initial value |
| $S$ | $(1 - e)(N - I_0)$ | 849999 | $S_d$ | $e(N - I_0)$ | 4249993 |
| $E_1$ | $0.4(1 - e)I_0$ | 0.53 | $E_{1d}$ | $0.4eI_0$ | 2.67 |
| $E_2$ | $0.1(1 - e)I_0$ | 0.13 | $E_{2d}$ | $0.1eI_0$ | 0.67 |
| $I$ | $0.5(1 - e)I_0$ | 0.67 | $I_d$ | $0.5eI_0$ | 3.33 |
| $Q$ | 0 | 0.00 | $Q_d$ | 0 | 0 |
| $R$ | 0 | 0.00 | $R_d$ | 0 | 0 |

### 2.4 Observation model

We used a Bayesian statistical model to condition our inference about *R*_0b_, *f*_2_, and expected case counts *µ*_*r*_ on the observed case count data *C*_*r*_. We related the expected number of cases to the observations through a negative binomial observation model using the NB2 parameterization of the probability mass function [28]:

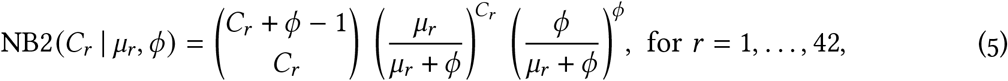

in which the variance scales quadratically with the mean according to the (inverse) dispersion parameter, *ϕ*: Var [***C***] = *µ*+*µ*^2^ /*ϕ*. Using the notation [*a* |*b*] to describe the conditional distribution of *a* given *b* and the bold symbol ***C*** to represent the vector of cases, we can write the joint posterior distribution for the model as

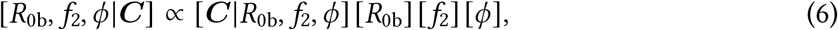

where the term [***C***| *R*_0b_, *f*_2_, *ϕ*] refers to the negative binomial data likelihood and the terms [*R*_0b_], [*f*_2_], and [*ϕ*] refer to the prior distributions.

We placed weakly informative priors on *R*_0b_, *f*_2_, and *ϕ*. We chose a prior on *R*_0b_ to encompass values commonly published in the literature for SARS-CoV-2 [2, 29]: Lognormal (log (2.6), 0.2). We chose a prior for *f*_2_ that resulted in a mean of 0.4 and a standard deviation of 0.2, *f*_2_ ∼Beta (2, 3), to represent a moderately strong reduction in contact fraction while still being broad enough to encompass a wide range of values. We chose a prior on *ϕ* to constrain the model to avoid substantial prior mass on a large amount of over-dispersion (small values of *ϕ*): 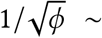 [30].

We fit our models with Stan 2.19.1 [31, 32] and R 3.6.2 [33]; Stan implements the No-U-Turn Hamiltonian Markov chain Monte Carlo algorithm [34] for Bayesian statistical inference. In our main model run, we sampled from eight chains with 2000 iterations each and discarded the first 1000 iterations of each chain as warm-up. We assessed chain convergence visually via trace plots (Figure S7) and via ensuring that 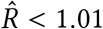 (the potential scale reduction factor) and that ESS *>* 200 (the effective sample size), as calculated by the rstan R package [32]. We represented uncertainty via quantile-based credible intervals.

### 2.5 Model validation

To test the model’s ability to recover the parameters and to test our implementation of the Stan model, we repeatedly fit the model to data simulated from separate code in R. We inspected the resulting posterior distributions for bias and coverage of the known true values and visually inspected the resulting time series (e.g., Figures S3 and S4). Furthermore, we conducted posterior predictive checks [35] to assess whether the observed data were consistent with data generated by the model. Code to reproduce our analysis is available at https://github.com/carolinecolijn/distancing-impact-covid19.

## 3 Results

We find that physical distancing has considerably reduced the contact rate. Our estimate of the fraction of contacts that have been removed due to distancing (1−*f*_2_) is 0.78 (0.66–0.89 90% CI [credible interval]), which is somewhat above the critical threshold (0.45; Figure 4). Figure 4 shows the case counts with the model fit, along with the posterior estimates for *R*_0b_ and the fraction of contacts that have been removed due to distancing measures. The model describes the count data well, with reported cases showing a peak in late March, approximately two weeks after the initiation of distancing measures (Figure 4A). The data are informative with respect to both main parameters, with the posteriors distinctly different and more peaked than the priors (Figure 4B,D). Figures S5–S7 show posterior samples for the model variables, examples of predicted case counts, and trace plots illustrating MCMC convergence. Figure S8 shows the joint posterior scatterplot. We have used a fixed value of *e*, the fraction engaged in distancing; this choice is motivated by the survey and behavioural data. If *e* were lower, the estimated strength of distancing would be higher to achieve the same case counts. We find this tradeoff analytically using the basic reproduction number for the full model (Supplement, Figure S1).

**Figure 4:**
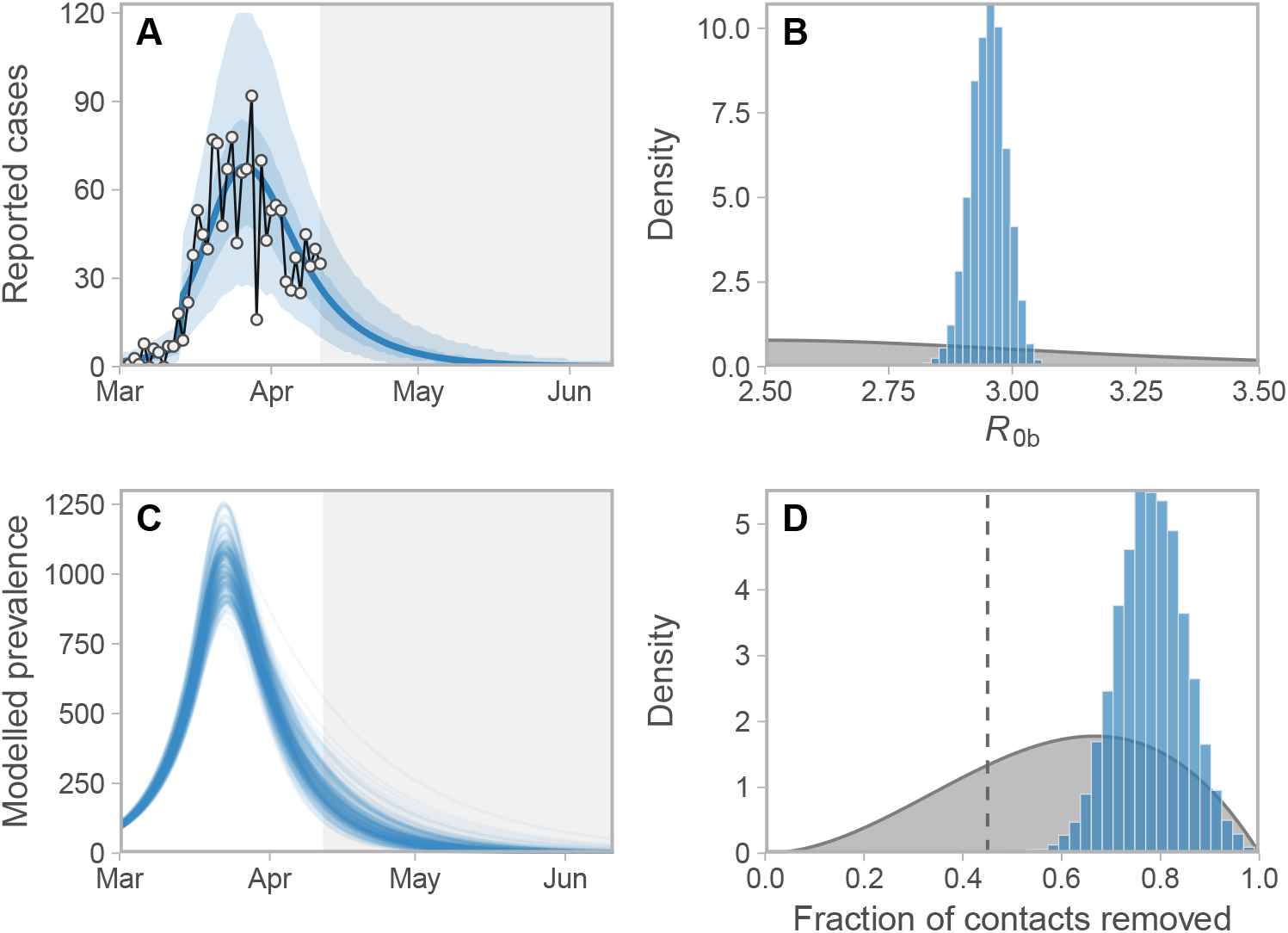
(A) Observed and estimated case counts, (C) estimated prevalence, and posterior estimates for (B) *R*_0b_ and (D) fraction of contacts removed (1− *f*_2_). These projections do not account for introduced cases from other jurisdictions and they assume that distancing measures remain in place. The fraction of contacts removed is the model’s portion of contacts that have been stopped among those who are engaged in physical distancing, with 0.0 meaning no change and 1.0 meaning 100% of contacts have been removed for those engaged in physical distancing. In panel A, the blue line represents the posterior mean and the shaded ribbons represent 50% and 90% credible intervals on new observations. Dots and black lines represent the reported data. In panel C, lines represent example draws from the posterior. In panels B and D, priors are shown in grey and posteriors in blue. In panel D, the dashed vertical line denotes the threshold below which an exponential increase in prevalence is expected (see Figure S9). Note: Model prevalence depends on our assumptions about underestimation, incubation period, and the duration of infection, none of which we can estimate well from these data (Figures S12). This model does not take asymptomatic individuals into account. Much higher values of the prevalence are consistent with our data.

We find that with a shorter incubation period and duration of infectiousness, a lower reproduction number will fit the same overall growth rate, and conversely if the infectious duration (and serial interval) are higher, a higher reproduction number is required but the fit to data is very similar (Figure S10). This relationship is well known [36]. The conclusion that distancing measures have reduced contact is robust to these alternatives (Figure S10). The model depends on the fraction engaged in distancing, but not much on the rates *u*_*d*_ and *u*_*r*_ ; Figure S11 illustrates that we obtain the same results with these rates increased by a factor of 10. We also explore the robustness to the unknown underestimation fractions (Figure S12), and a random walk pattern in the fraction of cases sampled (Figure S13); again, the data are consistent with a range of underestimation fractions, and the conclusion about the contact reduction is robust.

Our estimates suggest that some relaxation of current distancing measures would be possible without bringing the growth rate above zero (Figure 5A,D), but if measures are relaxed too much (in the absence of monitoring and re-starting measures), the prevalence and case counts begin to increase exponentially (Figure 5B–C,E–F), reaching high levels by June 2020 if distancing were to cease entirely (Figure 5F; Figure 6). These are illustrative scenarios only; public health responses with renewed or revised measures would likely be put in place rapidly were such rises to be observed. The speed of growth depends on how close the system is to the epidemic threshold (Figure S9). If strong enough measures are not maintained, the model predicts a range of possible epidemic curves (Figure 6) consistent with published simple and complex models [11, 10, 13].

**Figure 5:**
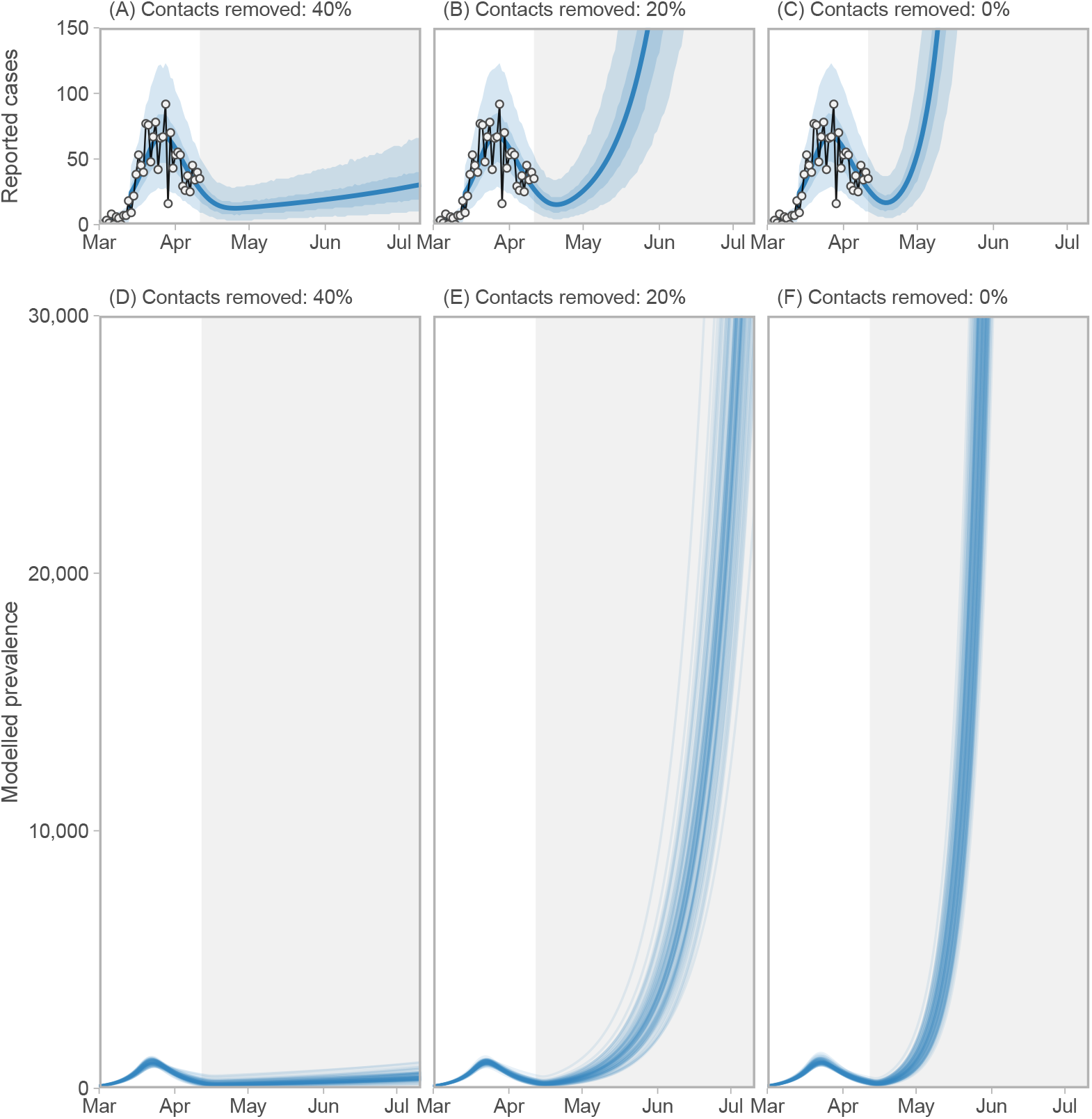
Three scenarios for relaxing distancing measures beginning on April 12 2020. (A, D) Distancing measures are relaxed but prevalence and reported cases continue to decline, with slower decline and more variance in the number of reported cases in coming weeks. (B–C, E–F) Distancing measures are relaxed (B, E) or cease (C, F) and exponential growth is observed beginning mid-April. Figure description is otherwise the same as for Figure 4A, C. Note: Model prevalence depends on our assumptions about underestimation, incubation period, and the duration of infection, none of which we can estimate well from these data (Figures S12). This model does not include individuals who do not develop symptoms. Much higher values of the prevalence are consistent with our data.

**Figure 6:**
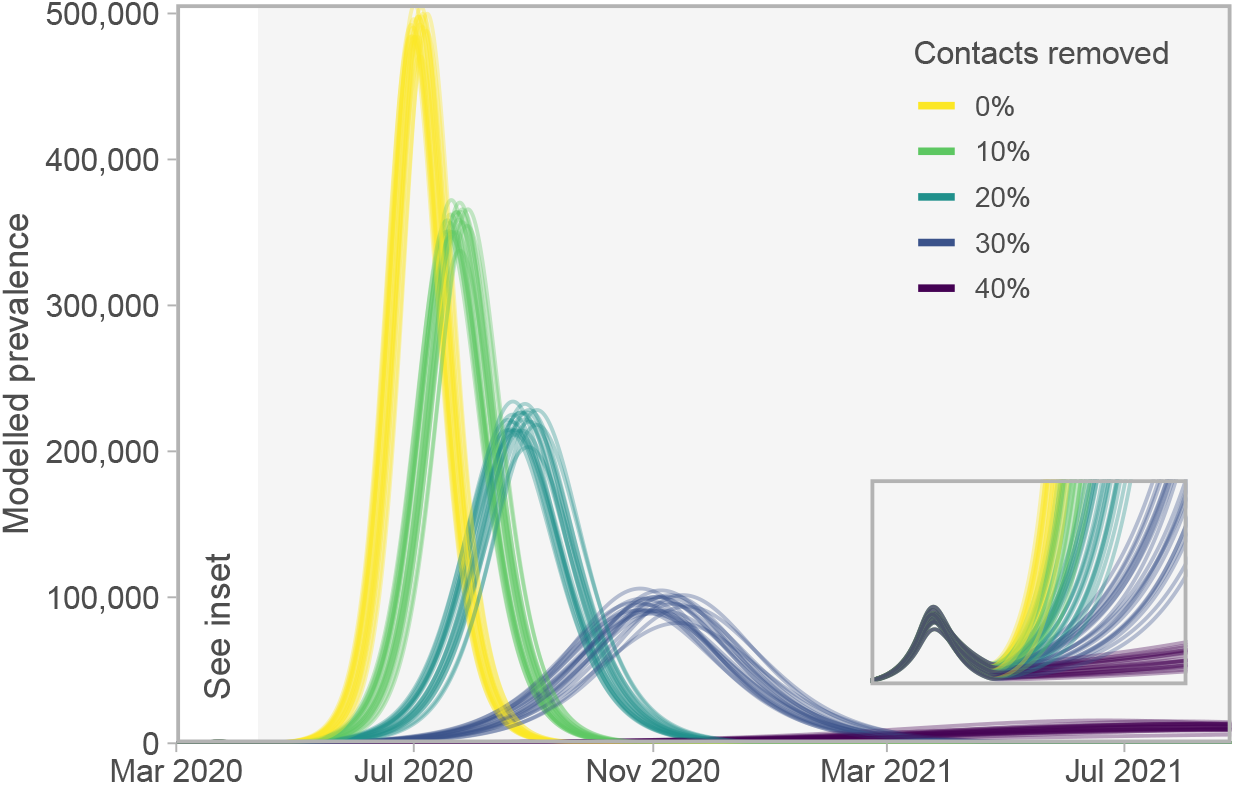
Epidemiological curves for five scenarios of relaxing distancing measures (beginning on April 12 2020) that are insufficient to keep the prevalence from growing. Stronger measures “flatten the curve”, as would be expected. Lines represent 20 draws from the joint posterior distribution. Grey region denotes future dates during which the relaxed distancing measures are applied. Inset box magnifies the lower-left region of the plot to show the modelled prevalence from March 1 to July 1 2020—the initial curve is imperceptible on the main axes. Note: Model prevalence depends on our assumptions about underestimation, incubation period, and the duration of infection, none of which we can estimate well from these data (Figures S12). This model does not include individuals who do not develop symptoms. Much higher values of the prevalence are consistent with our data.

There has been some interest in relaxing distancing measures and re-introducing them when a threshold has been reached, such as when intensive care capacity is nearly reached [37]. We did not explore a dynamic trigger, but did explore the behaviour when distancing measures are introduced and relaxed repeatedly (Figure 7). If the relaxation period is such that the outbreak remains contained throughout (with an effective reproduction number less than one), then the prevalence declines at alternating faster and slower rates. Simulations in which the relaxation of distancing measures allows prevalence to rise again are of more interest. In a scenario of switching between the current mean estimate (78% of contacts removed) and 20% contacts removed, reported cases rise, lagging the relaxation of distancing (Figure 7). Illustrative simulations in which distancing alternates every three or four weeks allows an overall continued decline; however, the longer the period of relaxation, the more the prevalence is able to rise in between periods of distancing (Figure 7). Control of delayed feedback systems is challenging [38], and ideally if a dynamic trigger such as reported or ICU cases were to be used, monitoring would need to be as rapid as possible. Monitoring of distancing behaviour and population contact patterns would be important, in addition to monitoring cases.

**Figure 7:**
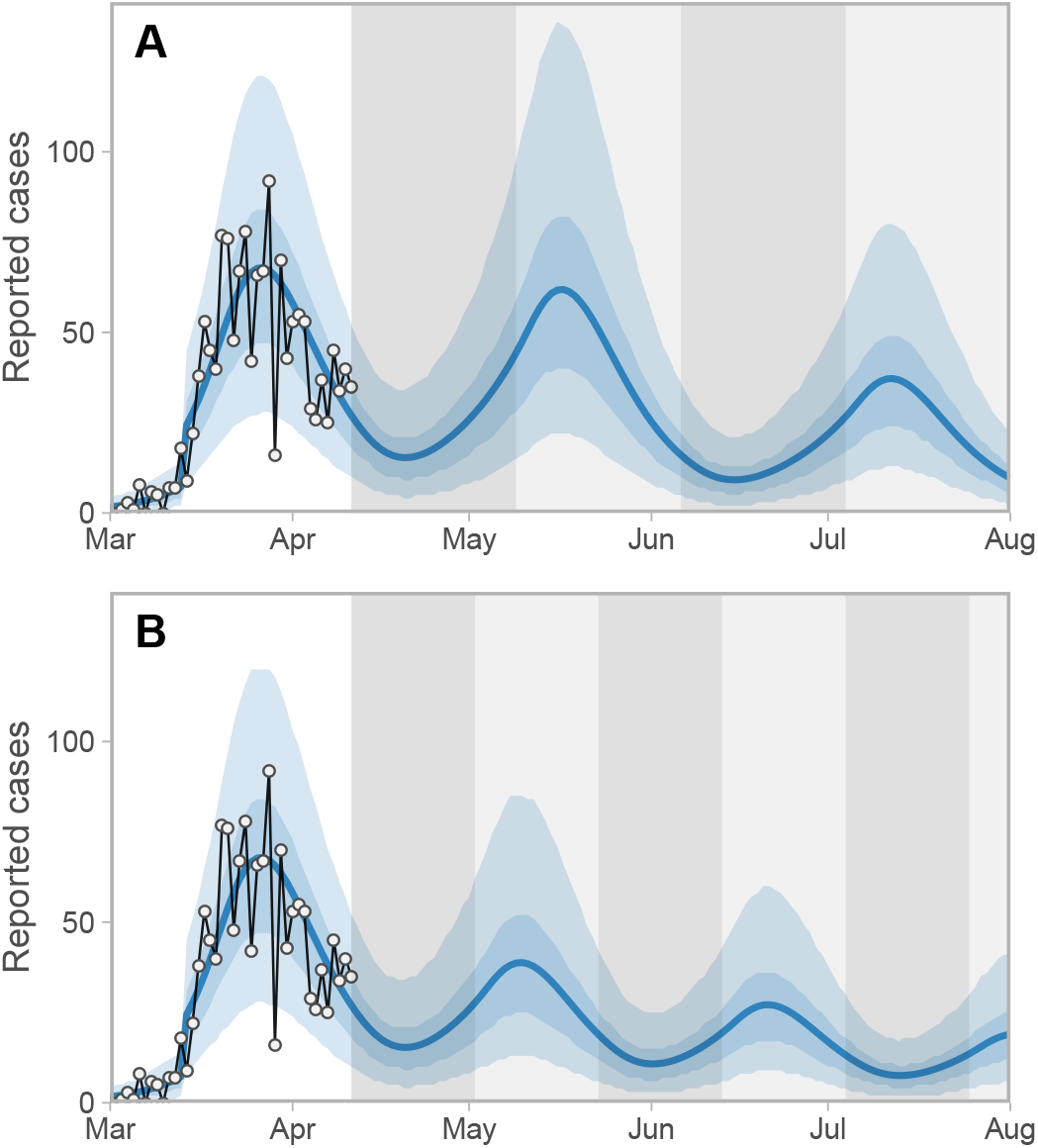
Two scenarios of cycling between physical distancing levels. Here, the percentage of contact reduction alternates between 20% and 78% at (A) four-week and (B) three-week intervals. Reducing contacts by 78% is approximately the estimated level by our model (Figure 4). Dark- and light-grey shaded regions represent the 20% and 78% time periods, respectively. Note the lag between changes in physical distancing and reported case counts. Blue lines represent the posterior mean and shaded ribbons represent 50% and 90% credible intervals on new observations. Dots and black lines represent the reported data.

## 4 Discussion

Our results suggest that physical distancing measures are effective in British Columbia at present. We estimate the fraction of contact reduction to be 0.78 (0.66–0.89 90% CI), which is stronger than the critical threshold of the model. Our conclusions about the decline in cases are supported by declines in hospitalizations and ICU admissions. Our estimate is in line with survey data from the UK [39] which suggested a 73% reduction in contacts, and a modelling study which found that a 70–80% reduction in contacts is consistent with data in France to date [12].

Our estimate of the effect of distancing on contact patterns is consistent with independent local lines of evidence for the strength of distancing measures. Local rail (Skytrain) station crowding data, provided by Metro Vancouver’s transportation authority TransLink, gives a proxy for reduction in public transport use. Between the weeks of February 24 and March 23, travel from 7–10am was reduced by 73% (on weekdays) and 63% (on weekends), from 11am–4pm by 71% and 72%, from 5pm–8pm by 79% and 80%, and from 8pm–12am by 73% and 76%. Overall daytime travel was reduced by 16% for the week of March 9, 64% for the week of March 16, and 73% for the week of March 23. Estimates on adhering to physical distancing are also available from a publicly available respondent-driven survey [18]. The survey found a the rate of respondents stating that there was a serious threat of a coronavirus outbreak in Canada increased from 42% on March 5–6 to 88% on March 20–23. For individuals who stated there was a serious threat, 89% stated they were keeping personal distance compared to 66% for individuals who did not believe there was a serious threat. Mobile phone location data from BlueDot [40] suggested that the maximum and cumulative distance travelled from home fell by approximately 90%, the portion of mobile phone check-ins at home rose by over 10%, and the portion of devices for which every check-in was at home rose by 60%. These estimates are also consistent with Google mobility [41], Citymapper index [42], and Apple mobility data [43]. These are indirect reflections of the contact rate but are supportive of a dramatic change in contact patterns as reflected in our model.

Our results suggest that some relaxation of distancing measures may be possible; we have simulated fixed and dynamic measures, less stringent than the measures in place at present, that would continue to maintain low case numbers. This is feasible either through continual distancing that is quite strong, or via well-monitored dynamic on/off measures. We have illustrated the model’s high case volumes and long time frames that would result from cessation of distancing and the absence of continued strong public health and behavioural intervention. These simulations illustrate that immunity has not built up in the model; our estimates of the decline are not due to a natural peak in an epidemic curve, but are the direct effect of changes in the contact patterns in the population. Seasonal transmission could even amplify a peak in the winter season, if control and monitoring were ceased then [11]. However, experience to date indicates that given any early signs of renewed growth, public health in BC is in a position to manage and reduce transmission, as evidenced by current declines in BC; the benefit of recent experience could enable the response to be swifter and more effective if case numbers begin to rise again.

Distancing measures were in place in BC from approximately March 15 and we believe that they had taken full effect by the week of 22 March. The peak in observed cases occurred at approximately March 27, 12 days after the model’s ramp-up of distancing began and 15 days after the initiation of any distancing measures. This delay presents obvious challenges for monitoring the success of control measures using reported case count data. We suggest that two different kinds of monitoring will be important if distancing measures are to be relaxed: (1) monitoring cases, through testing, contact tracing, and other case finding, and (2) monitoring contact patterns and distancing behaviour in the population, in a kind of “distancing surveillance” effort. These data, derived from mobile phones, surveys and apps, could be available very rapidly, whereas the incubation period places an unavoidable delay between control measures and detecting their impact in reported cases– even if testing were widespread and reporting were immediate. There is considerable interest in real-time monitoring of mobile phone movements, population surveys on the uptake of physical distancing, and other behavioural data. While these are potentially promising avenues, the outcome of interest is incident infections. Locations of mobile phones, traffic patterns, and survey information are all proxies for this outcome at best. The work we have presented here could help to calibrate distancing surveillance measurements, to understand how they relate to changes in contact rates for modelling efforts.

Our modelling framework has a number of important limitations. We do not model age and contact structure explicitly, except to distinguish between two populations: those participating in distancing or not. This has the advantage that we do not require data on age-specific contact patterns, responses to distancing measures, or infectiousness; these data are not available at this time. It also limits our ability to provide guidance on where and how contact reduction measures could be implemented. It is a simplification of behaviour in many ways; true distancing responses are a continuum, and the measures in place (e.g. no mass gatherings or dine-in services) also mean that the whole population is experiencing some changes in contact patterns. Our model is deterministic, and so does not capture the possibility of extinction; in addition, we have not simulated introductions of COVID-19 from other jurisdictions. We have not accounted for geographic structure; differences in distancing behaviour, health care practices, and demographics in different jurisdictions could impact the results. We have not modelled either conventional or automated contact tracing [44, 45]; in our model these would decrease the duration of the infectious period and change the transitions for some exposed individuals.

There are also limitations in our data. We have used an observation model to link reported cases to the modelled prevalence, and we included variation in the portion of cases detected over time. Modelling and forecasting based on reported cases faces challenges when testing is driven by clinical needs, testing capacities, and other constraints (and in particular is not designed to test population samples). Cases in long-term care facilities (LTCF) represent a substantial fraction of the cases, and particularly the deaths, in BC. Along with the low number of deaths in total, this is one rationale for not modelling deaths explicitly. We included LTCF cases but also modelled a wide range of under-reporting to account for potential biases. If many cases in an LTCF cluster were all reported on the same day (or within a short time frame) this could increase the noise in reported case counts. We have modelled case counts as over-dispersed to account for such variation.

The testing criteria include a number of categories that have changed over time. Testing volume increased sharply in mid March. In the latter half of March and until April 9, testing focused primarily on those hospitalized or likely to be hospitalized, health care workers, residents of long-term care facilities, and other cluster investigations. After April 9, testing was opened somewhat to include residents of remote, isolated or Indigenous communities, people who are homeless or have unstable housing, and by physicians’ clinical judgement. There is likely some inconsistency in the application of these guidelines across hospitals and facilities. As a consequence, the base population being tested changes with time and contributes a changing portion of the force of infection. Underestimation is therefore complex and is comprised of varying under-ascertainment and under-reporting.

There remain important unknowns about COVID-19 that give rise to additional limitations for modelling efforts; immunity and asymptomatic transmission are two of these. We have included pre-symptomatic transmission but we have not explicitly modelled asymptomatic individuals, who may have few or no symptoms but nonetheless be transmitting, and who may or may not be building lasting immunity. Recent work suggests that both pre- and asymptomatic individuals may be contributing considerably to transmission [46, 25, 26]. We have indirectly approached this uncertainty, exploring variable underestimation fractions and duration of the incubation and infectious periods. A wide range for underestimation and duration is consistent with the reported case data. Our conclusion about the impact of distancing measures appears to be robust to these uncertainties, although the basic reproductive number and the model prevalence vary according to which assumptions are made about underestimation and duration. Model predictions for the peak timing and size without strong public health interventions will depend strongly on the dynamics of immunity, including the numbers of asymptomatic individuals and their immunity [11].

Our model suggests that distancing measures are working in British Columbia, that some relaxation of these measures may be possible, and that this must be done carefully. Given the likely low levels of immunity, long-term public health measures will be necessary to control COVID-19. If data were available describing contact patterns and transmissibility by age, along with data describing the impact of specific activities on these contact patterns, then models could be effective tools to determine how to safely relax distancing measures—for example, restarting specific activities such as particular workplaces or schools. Without knowledge of prevalence and transmissibility in children, of the extent of asymptomatic infection, and of the contact patterns that would result from restarting specific activities, models aiming to simulate these activities will have large uncertainties. We therefore suggest that there is an urgent need for longitudinal measurements of population-level prevalence and immunity via viral testing and serological studies, even where prevalence is low, and despite this being challenging. This will inform our understanding of the role of children and schools. Furthermore, if distancing measures are to be relaxed, it will be crucial to have strong surveillance in place through widespread testing, contact tracing, and isolation of new cases, as well as strong compliance with potentially shifting public health policy and messaging.

## Data Availability

Data are available publicly at links given in the manuscript. The analysis is reproducible with code at the github link below.

https://github.com/carolinecolijn/distancing-impact-covid19

## Aknowledgements

We thank TransLink for providing data on transportation and Bluedot for descriptive summary statistics on mobile-phone mobility patterns. The research and analysis are based on data from TransLink and the opinions expressed do not represent the views of TransLink. The opinions and conclusions expressed also do not represent the views of Bluedot. We thank J.A. Otte for providing details on testing-criteria changes in BC and for insightful comments on an earlier version of this manuscript. C.C. and J.S. are funded by the Federal Government of Canada’s Canada 150 Research Chair program. We thank Fisheries and Oceans Canada for their support.

## A Supplemental Methods

### A.1 Computing *R*_0_ analytically

We compute the basic reproduction number *R*_0_ for the full model, as a function of the model parameters, using the Next-Generation method [47, 48]. We set *f* to be constant, dropping the subscript 2, and set *u*_d_ = 1, *e* := *u*_d_ / (*u*_*r*_+ *u*_d_), and *u*_*r*_ = (1− *e*) /*e*. The disease-free equilibrium is ϵ_0_ = ((1− *e*) *N*, 0, 0, 0, 0, 0, *eN*, 0, 0, 0, 0, 0). We identify *E*_1_, *E*_2_, *I, Q, E*_1d_, *E*_2d_, *I*_d_, *Q*_d_ as the infection compartments, and denote the corresponding system by *X*.

We decompose *X* = *F*− *V*, where

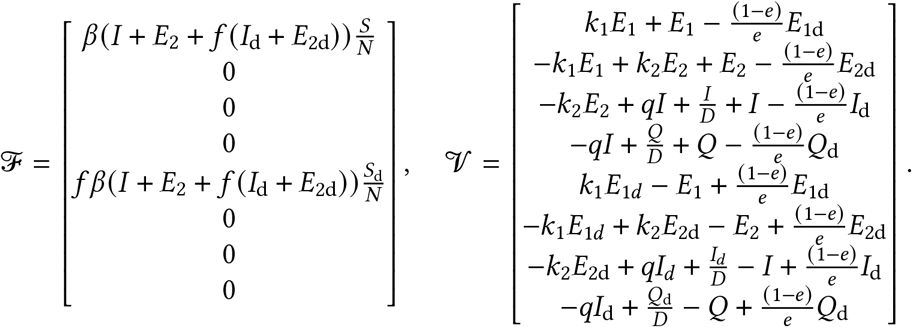

Linearization around ϵ_0_ gives the following Jacobian matrices for *F*and *V* respectively,

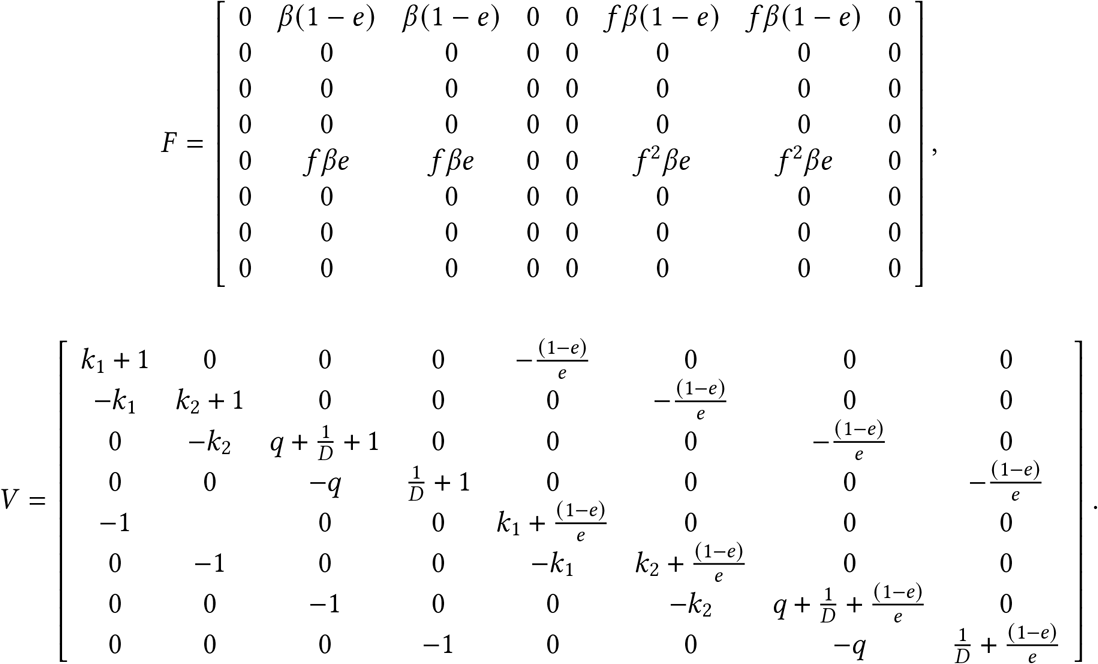

We use SageMath [49] via CoCalc [50] to compute the spectral radius of *FV* ^−1^ and simplify its expression. We find that

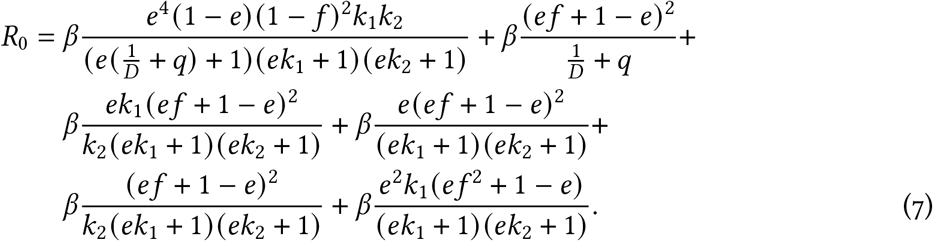

**Figure S1:**
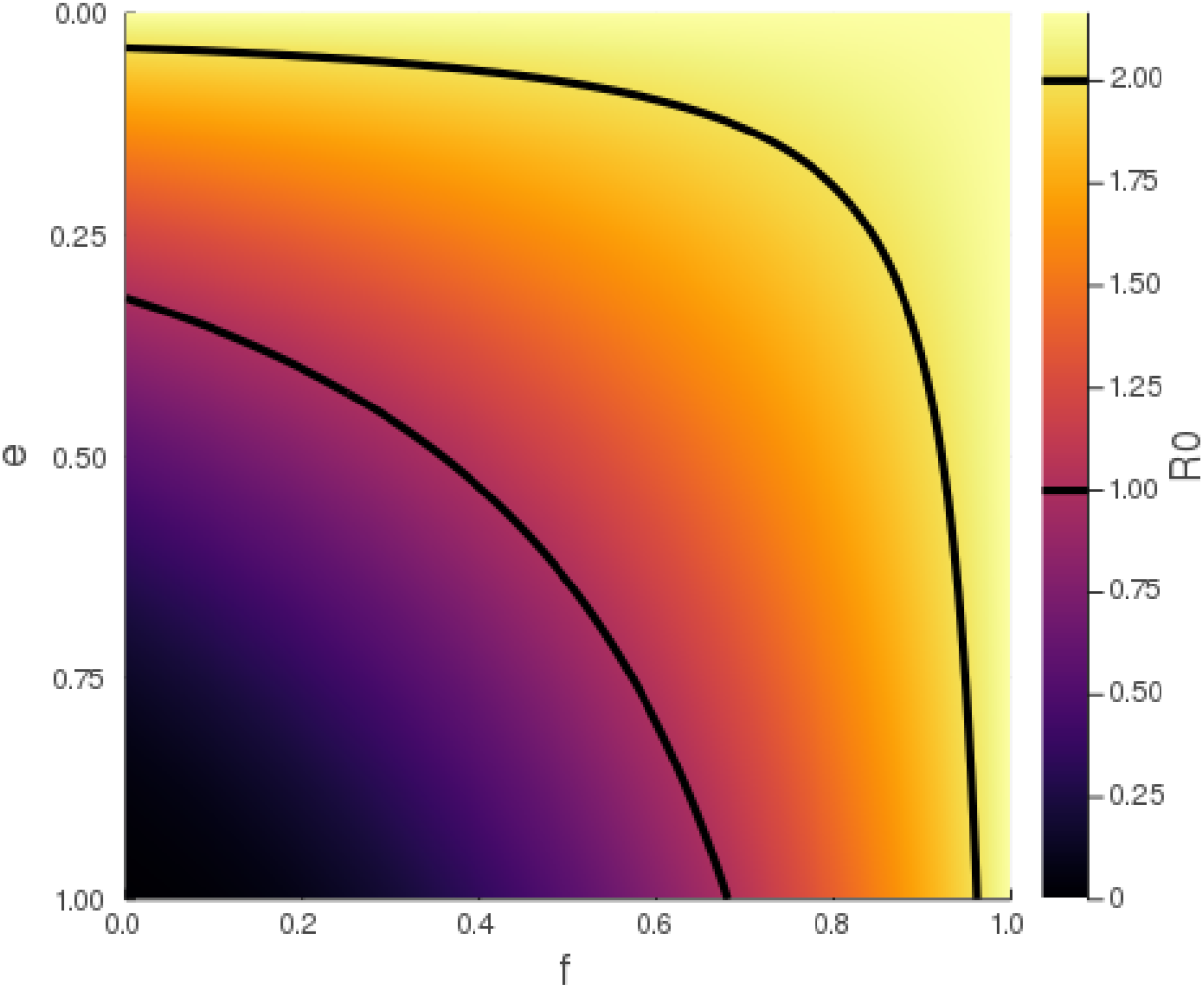
*R*_0_ as a function of *e* and *f*, with fixed parameters *β* = 0.433, *D* = 5, *q* = 0.05, *k*_1_ = 0.2, and *k*_2_ = 1. The black contours represent *R*_0_ = 1 and *R*_0_ = 2, respectively.

Figure S1 shows an example of the relation between *R*_0_ and the model parameters. The fixed parameters are taken from Table 1, with *β* computed as *R*_0b_/(*D* + 1/*k*_2_).

### A.2 Likelihood estimation of parameters for the delay distribution

We derive a likelihood function to estimate the shape and scale parameters of the Weibull delay distribution using the case-specific data that are available for some individual cases. These data are right-truncated (Figure 2), in that we are not aware of cases that will be reported in the future, yet their symptoms have already occurred in the past. This is dealt with by the likelihood function. The resulting maximum likelihood estimates are then used in the observation model to relate reported cases to model prevalence.

The data on individual cases (Figure 2) give counts *h*_*nr*_ of the number of individuals whose case was reported (test was positive) at the end of day *r* and whose symptoms are estimated to have started on day *n*. If day is the final day of these data, then the full data set is {*h*_*nr*_} _*n*=0,1,2,…, ;*n* ≤*r* ≤*N*_, where, by definition, *h*_*nr*_ = 0 for *r* < *n* because a case cannot be reported before the start of symptoms for these data. The counts are right-truncated on day, since there are individuals whose symptoms started on day *n* who will be reported in the future (after), but we do not yet know when. The cases are considered to be reported at the end of day *r* because there are values of *h*_*nn*_ *>* 0.

Assuming a Weibull distribution for the time between symptom onset and reporting of a case, then the probability of a case whose symptoms start on day *n* being reported on day *r* ≥ *n* is

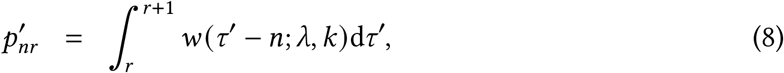

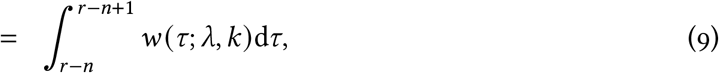

where the probability density function for the Weibull distribution with shape *k >* 0 and scale *λ >* 0 is

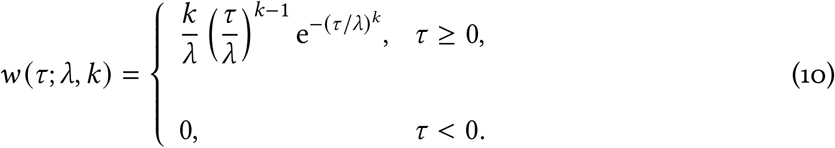

The integral in (9) is needed because the Weibull is a continuous function of time (and time is continuous in the differential equation model). We define the cumulative distribution function as

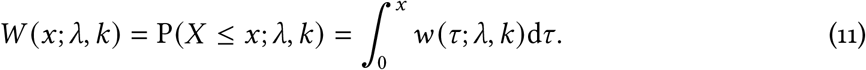

Then

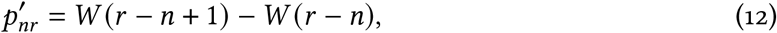

where we have dropped the explicit *λ* and *k* in *W*(·) for clarity.

Equation (9) holds for *r >N*, but the data are only available up to *r* =*N*. Consider that today is day *N* = 5, then we have a matrix of 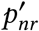 values:

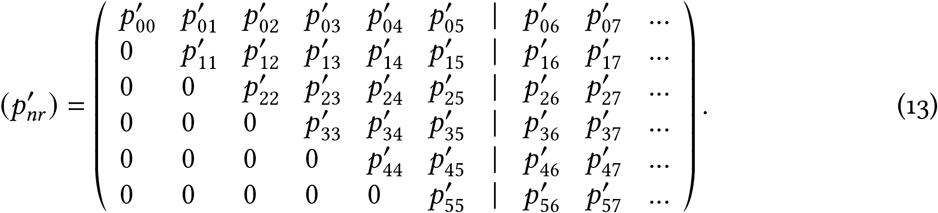

The probabilities in each row sum to 1 (since every case will eventually become reported, else it would not become a ‘case’). The cases to the right of the vertical dashed line are the ones that we do not yet have data on; for example, people whose symptoms started on day 1 and whose case will be reported tomorrow (day 6, i.e.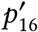).

We need to calculate *p*_*nr*_, the probability that a case that is reported on day *r* exhibited the onset of symptoms on day *n*. This requires taking the values in the above matrix that are to the left of the dashed line, and normalising each column such that it sums to 1, i.e.

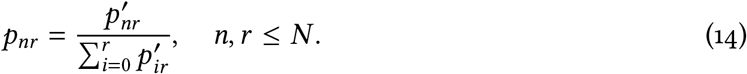

The denominator is

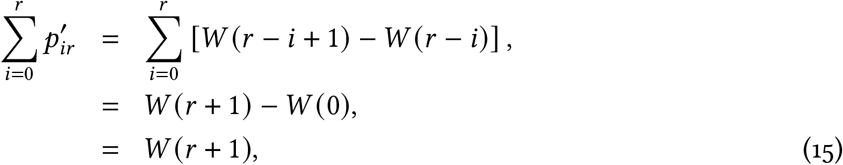

such that

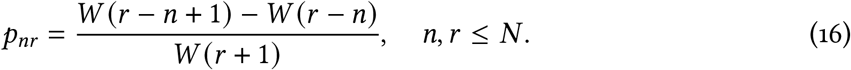

For = 5 we have the matrix of 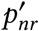 values:

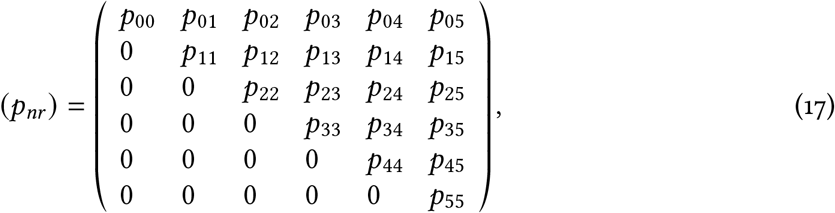

which is an upper triangular matrix of size 6 ×6, i.e. (*N*+1) × (*N*+ 1). With an extra day of data (setting = 6) the matrix has an extra row (people with onset of symptoms on day 6 can now be reported) and an extra column (the reported values on day 6):

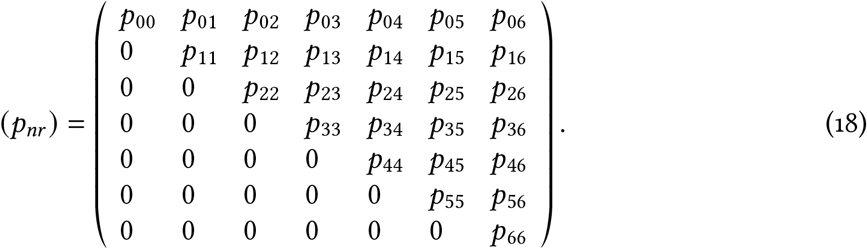

Given the data *h*_*nr*_ and these probabilities, we develop a multinomial log-likelihood function [19], adapting the approach of [20], to estimate the parameters *λ* and *k*. The log-likelihood function for *λ* and *k*, given the counts {*h*_*nr*_}, is

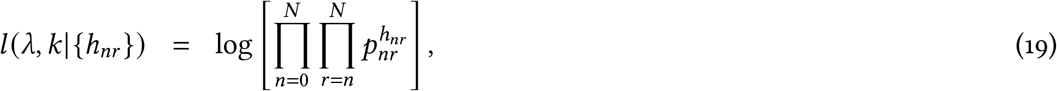

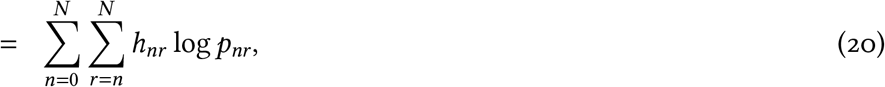

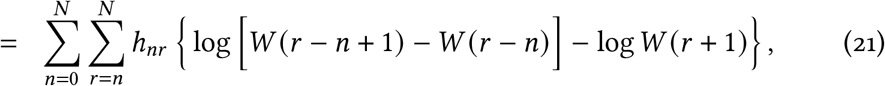

where the *W*(·) terms depend on *λ* and *k*. This is maximised numerically to give maximum likelihood estimates *λ*_MLE_ and *k*_MLE_ that are used in the differential equation model.

A key point to realise is that the definitions of *p*_*nr*_ for *r* = 0 to 5 (columns 1–6) are the same in (17) and (18), but their values will change from (17) to (18) because fitting the extra data from day 6 will update *λ*_MLE_ and *k*_MLE_, which affects all the estimated values of the probabilities. Also, not all of the daily reported cases used in the differential equation model have details regarding onset of symptoms, i.e.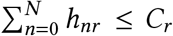. Using the available data we obtain estimates (with 95% univariate confidence intervals) of *λ*_MLE_ = 9.85 (9.30 −10.46) and *k*_MLE_ = 1.73 (1.60 −1.86), as shown in Figure S2. Using the maximum likelihood estimates, the mean delay is 8.78 days.

In the differential equation model, time is continuous and defined such that *C*_*r*_, the number of recorded cases on day *r*, is equal to the number of cases recorded by the end of day *r*. The number of people per day who become symptomatic throughout day *n* is the total number moving from the *E*_2_ and *E*_2d_ compartments to the *I* and *I*_d_ compartments, which is

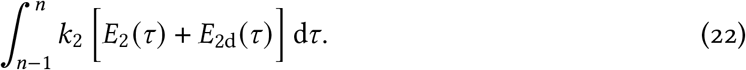

The expected number of cases that are reported on day *r* is then

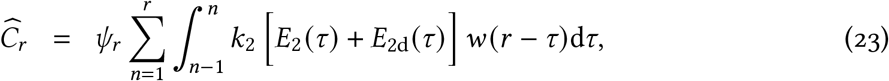

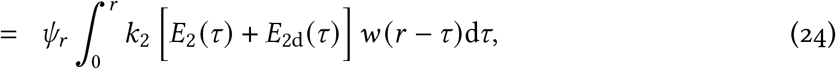

where *ψ*_*r*_ represents the sampling fraction on day *r* and *w*(·) represents the Weibull distribution with parameters *λ*_MLE_ and *k*_MLE_. If *ψ*_*r*_ = 1 then all estimated infectious people are tested and then become reported cases; *ψ*_*r*_ < 1 represents a reduction in expected cases on day *r* due to not everyone being tested. By changing variables *τ* = *r* − *s*, this becomes

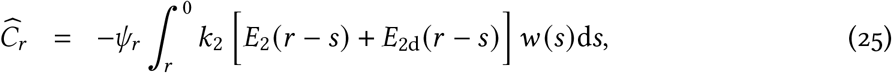

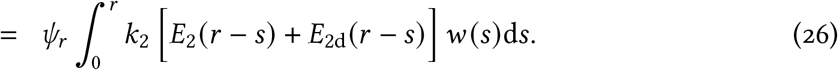

The differential equation model is started from a time earlier than *r* = 0, so the integration here can go back further because the model calculates, for example, *E*_2_ (−1); *r* < 0 does not occur in the above likelihood calculations because, by definition, there are no data for such times. This requires setting some maximum delay between symptom onset and a case being reported that is large enough for *w* (*s*) to be negligible. Based on Figure S2 we use a maximum delay of 45 days (the 99.99992% quantile), giving

**Figure S2:**
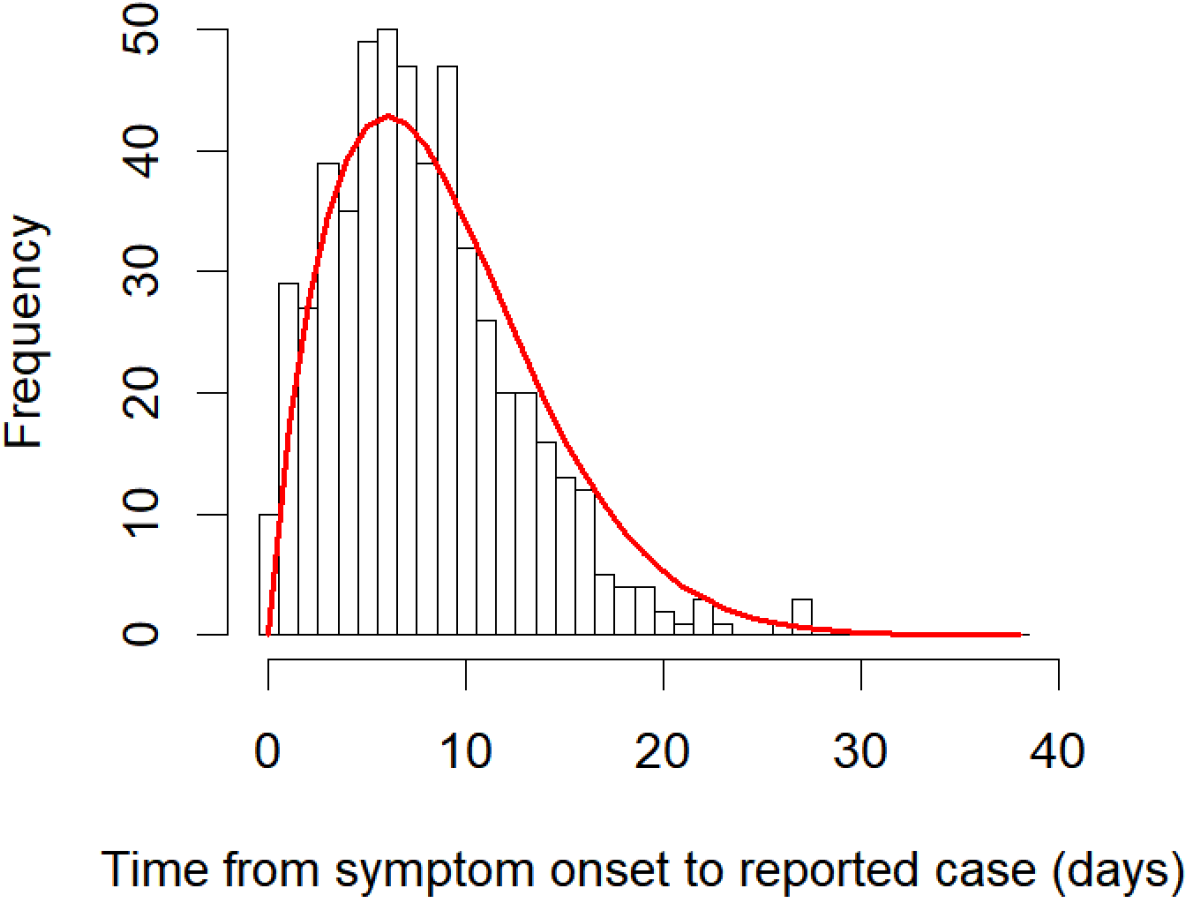
Histogram of times between onset of symptoms and a case being reported (from the case-specific data). The red line shows the maximum likelihood fit of the Weibull distribution, taking into account the right-truncation of the data. The distribution estimates longer delays (mean of 8.78 days) than shown in the data (mean of 7.79 days) due to the fit accounting for right-truncation—there has not yet been the opportunity to record many long delays (Figure 2).

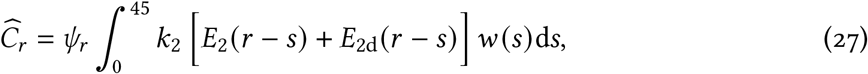

which is calculated and compared to the data (*C*_*r*_) via the negative binomial observation model.

### A.3 Model Validation

**Figure S3:**
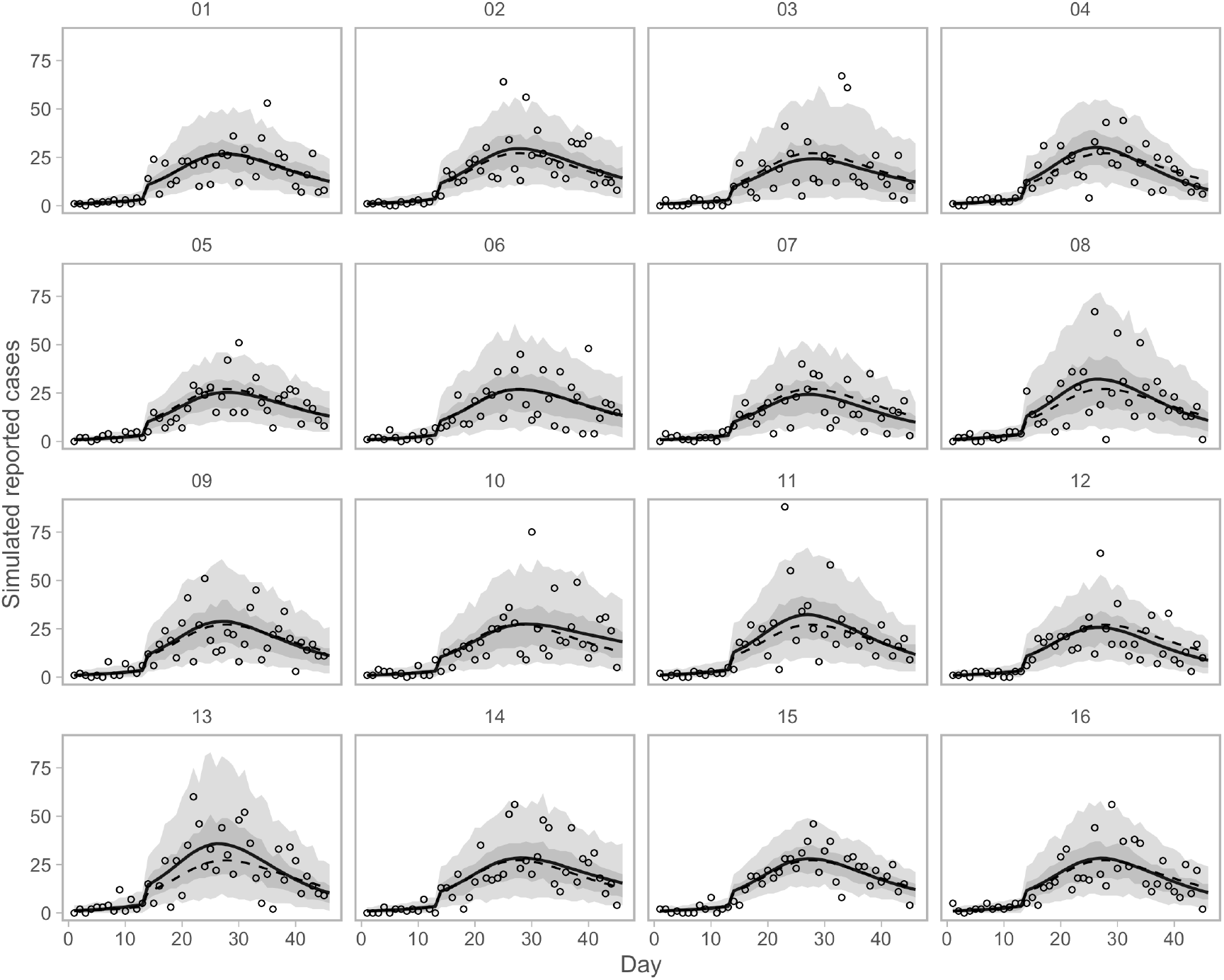
Simulation testing the ability to recover time series of reported cases. Dashed lines represent the true simulated time series. Solid lines represent means of the posterior. Dots represent observed simulated data. Shaded ribbons represent 50% and 90% credible intervals on new observations and should ideally encompass 50% and 90% of the simulated data points, respectively.

**Figure S4:**
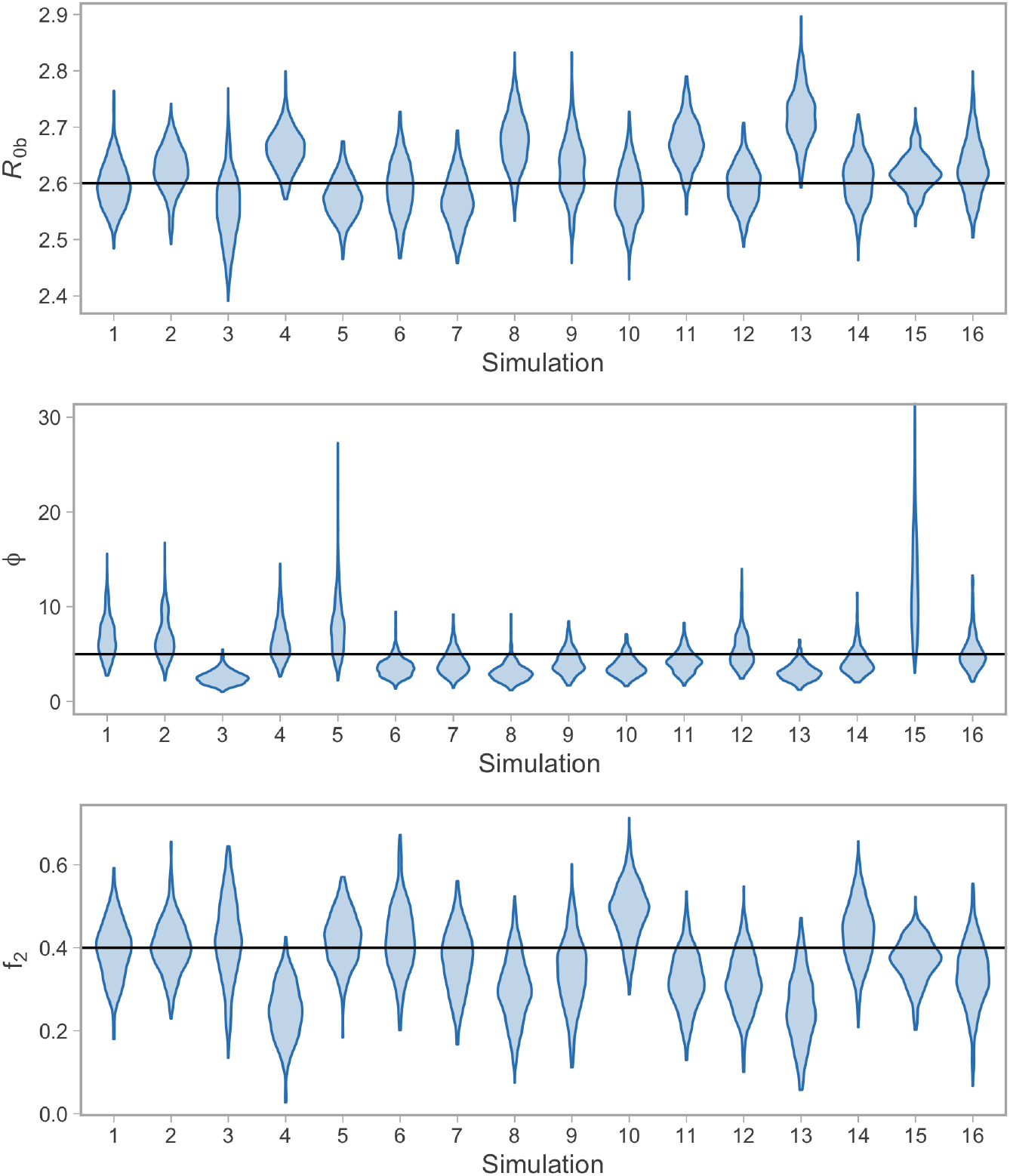
Simulation testing the ability to recover parameter values. The parameter *R*_0b_ is the basic reproductive number of the model without interventions. The parameter *ϕ* is the dispersion parameter of the negative binomial observation model. The fraction of contacts that are removed is 1− *f*_2_. The “violins” illustrate the posterior density distribution across 16 simulation examples. The true values used in the simulation are indicated by the horizontal black lines.

## B Supplemental Results

**Figure S5:**
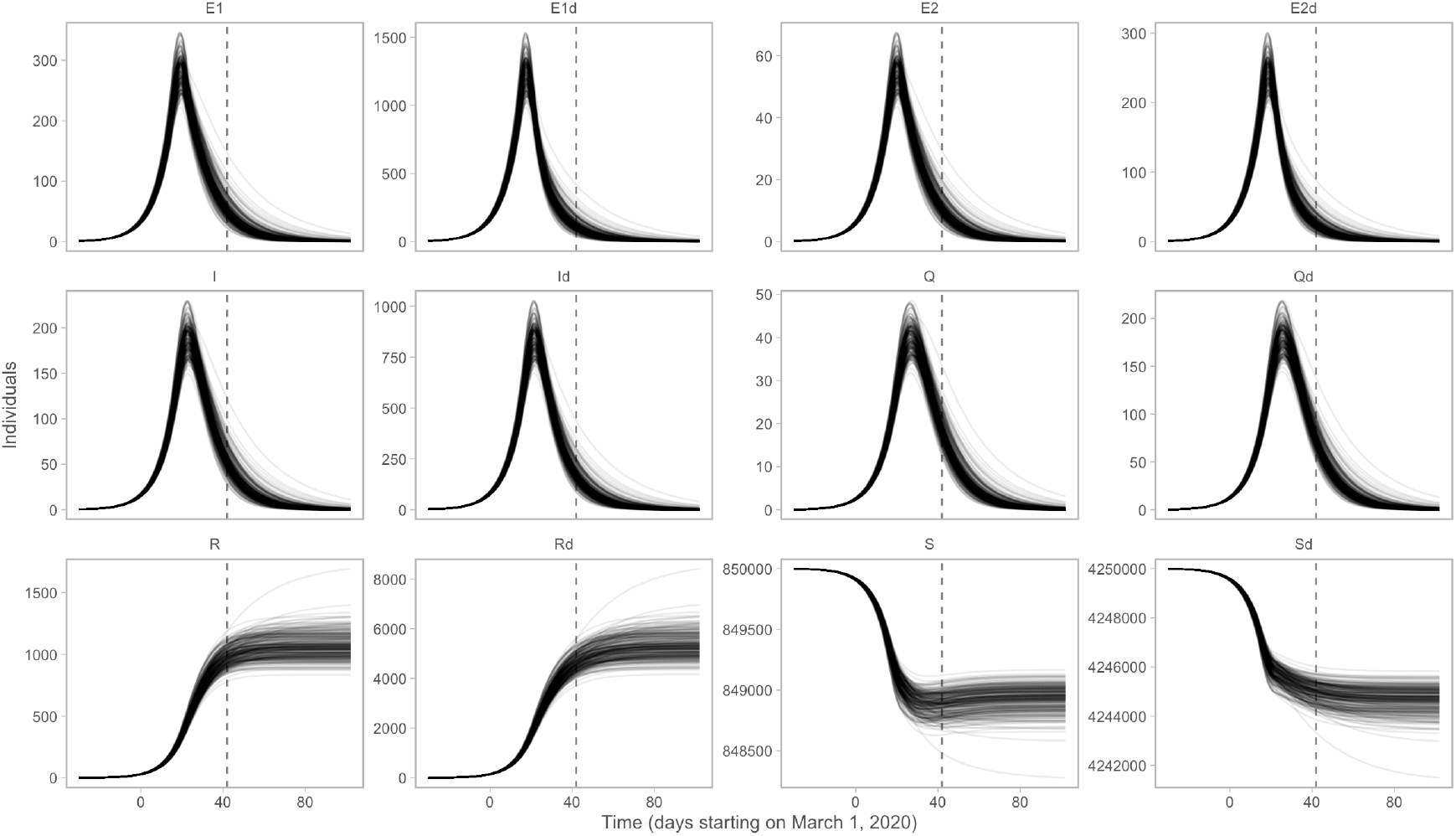
Posterior distributions of the underlying state variables. The time-dependent variables are the numbers of individuals who are: susceptible (*5*), exposed to the virus, asymptomatic and not infectious (*E*_1_), exposed, asymptomatic and infectious (*E*_2_), infectious (*I*), quarantined (*Q*), and recovered and deceased (*R*). Recovered individuals are assumed to be immune. There are analogous variables for individuals practising physical distancing, represented by subscript d, i.e., *5*_d_, *E*_1d_, *E*_2d_, *I*_d_, *Q*_d_, and *R*_d_.

**Figure S6:**
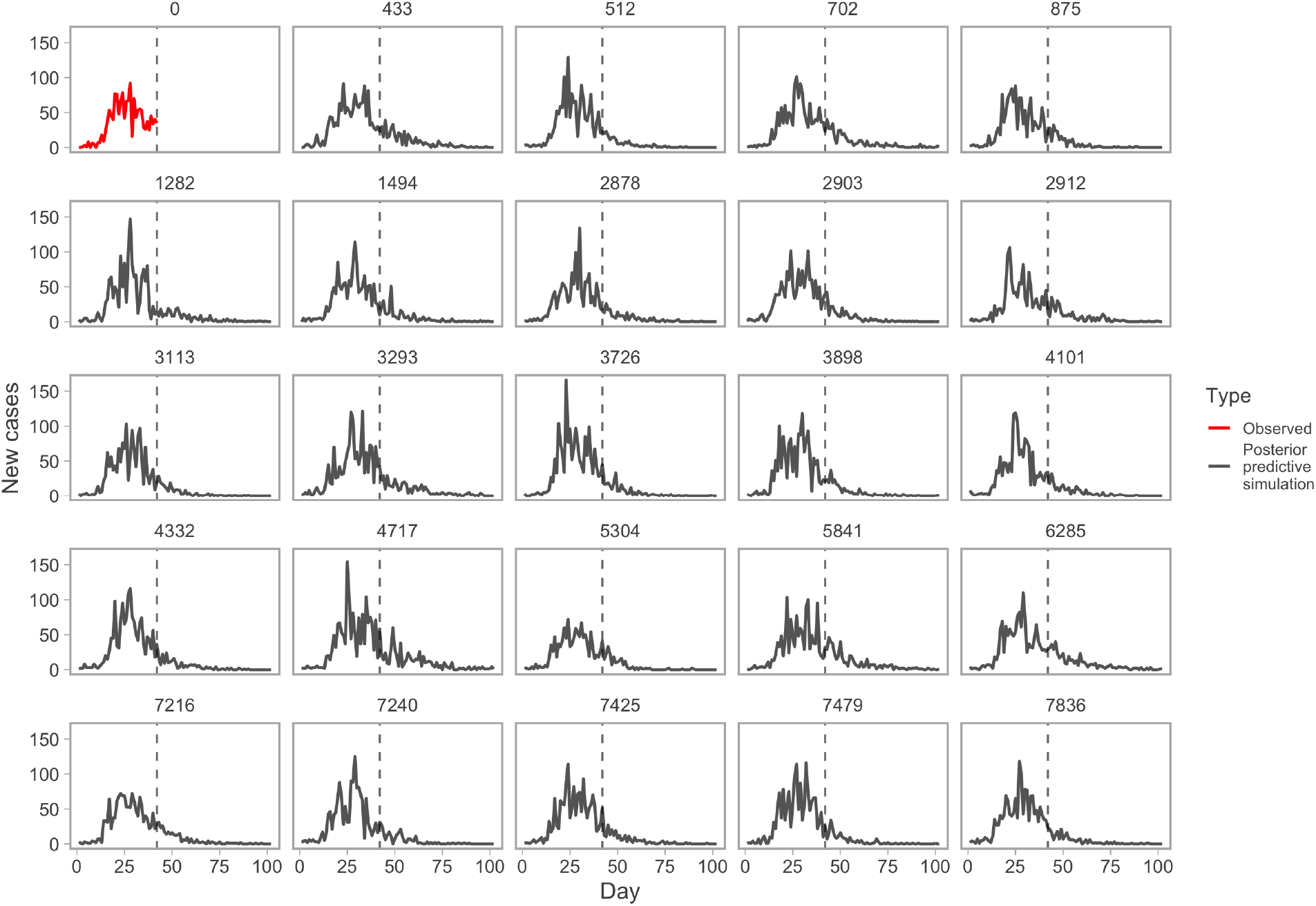
Example posterior-predictive replicates from the model fit. The first panel (red line) represents the observed data. All other panels represent example draws from the posterior-predictive distribution. Numbers above panels represent randomly selected Markov chain Monte Carlo (MCMC) iterations. This plot helps evaluate whether the observed data are consistent with data generated by the model.

**Figure S7:**
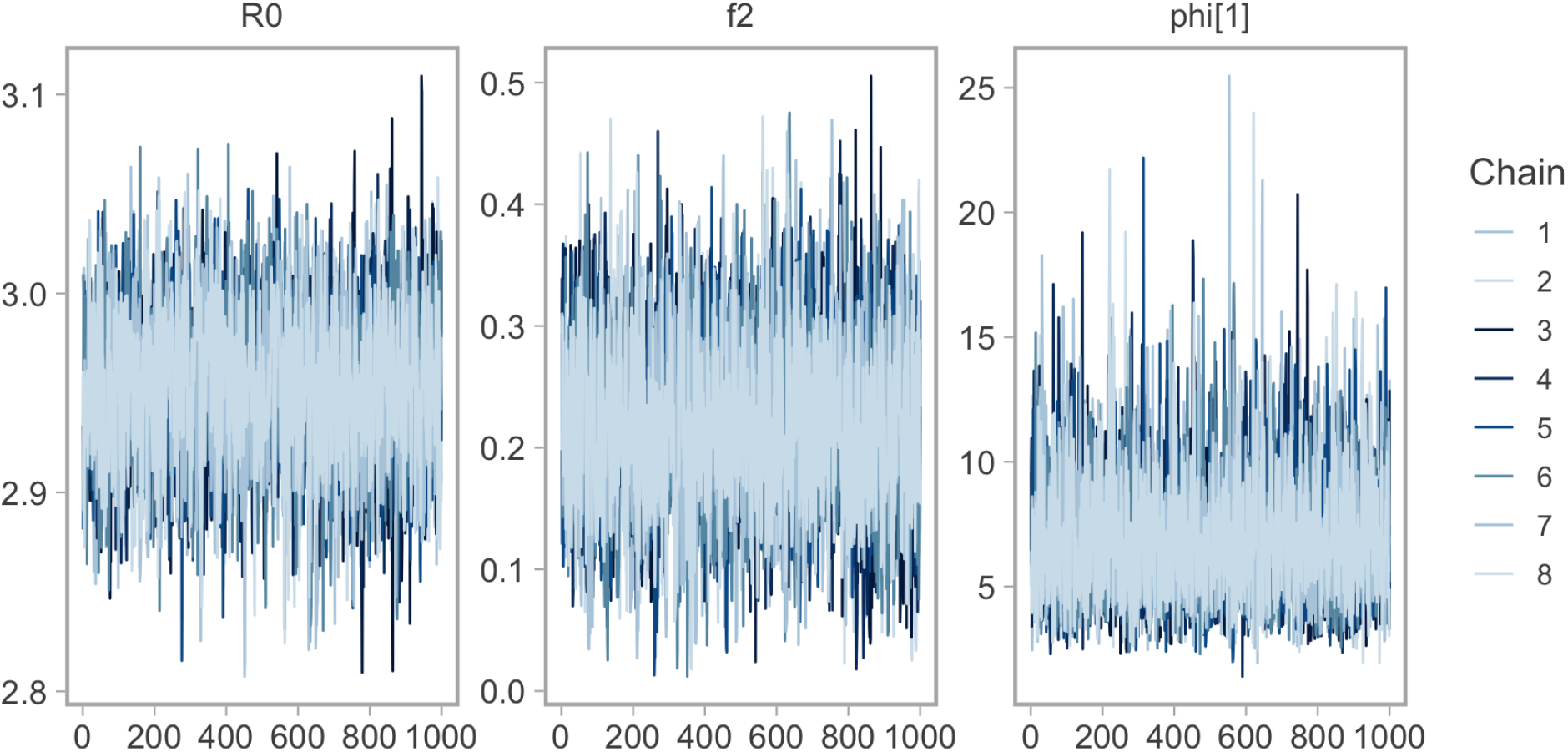
Trace plots of MCMC (Markov chain Monte Carlo) samples from parameter distributions to assess chain convergence. The panel labelled “R0” actually refers to *R*_0b_. The panel labelled “phi[1]” refers to the dispersion parameter *ϕ* from the negative binomial observation model.

**Figure S8:**
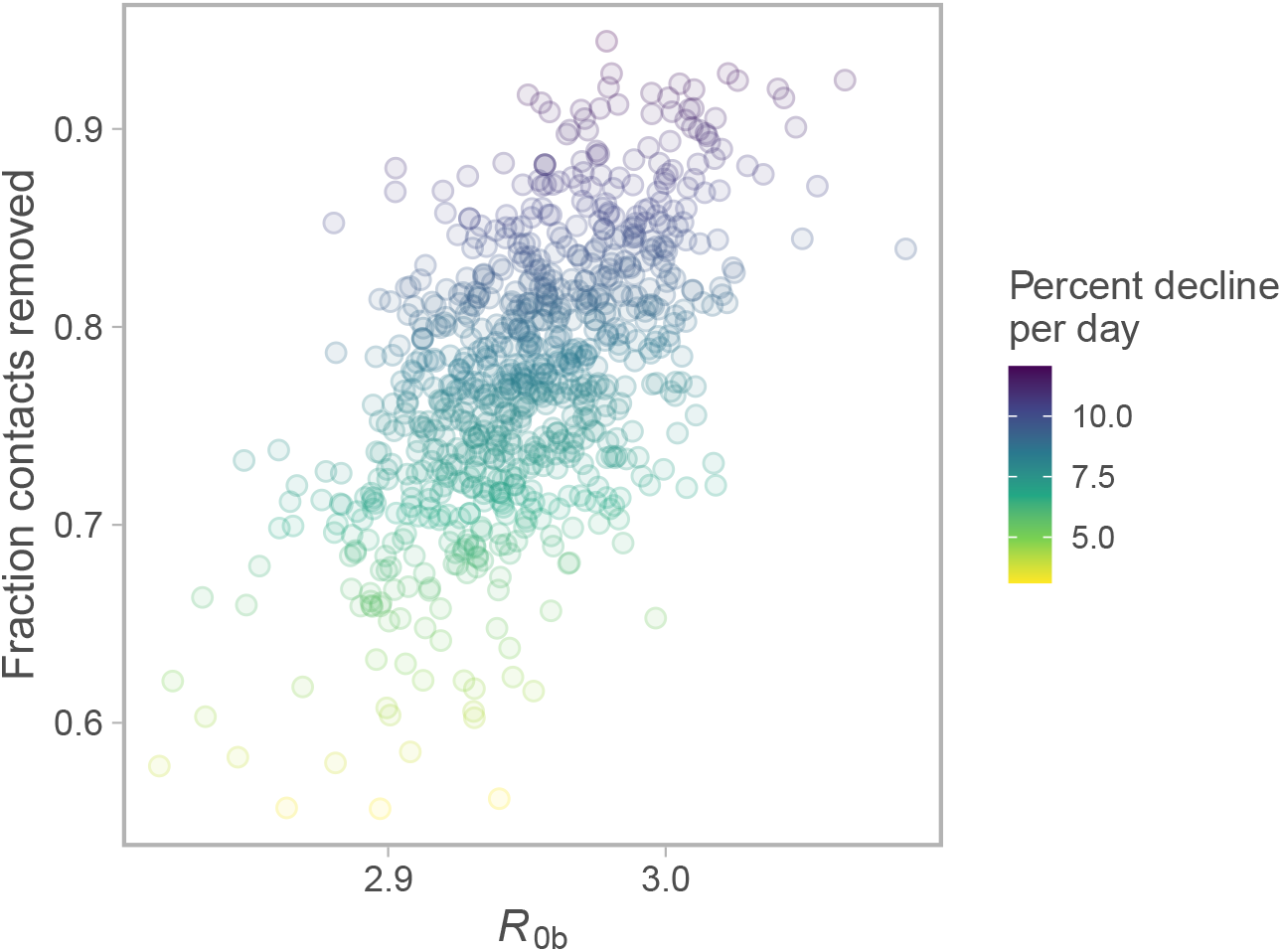
Joint posterior of fraction contacts removed vs. *R*_0b_. Colour represents percent decline per day over the last 30 days of the projection. Dots represent individual draws from the joint posterior.

**Figure S9:**
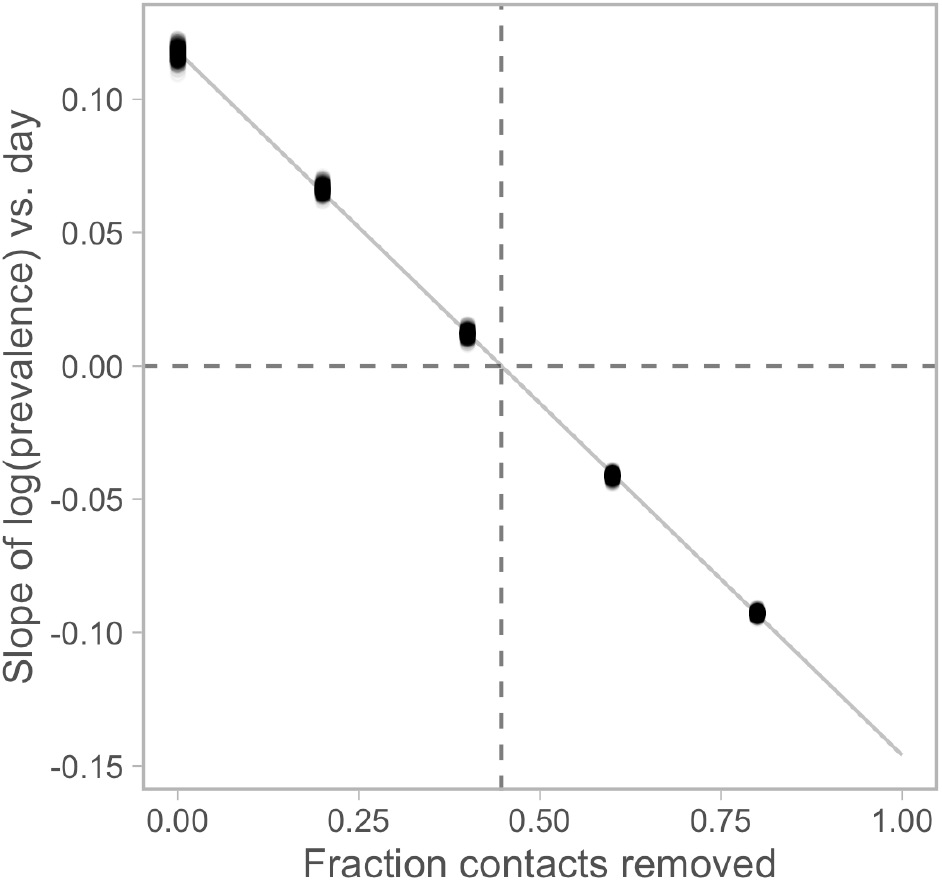
Linear regression of [the slope of log(prevalence) over days (for the last 30 days of the projection)] vs. [fraction of contacts removed]. Vertical dashed line indicates the critical value below which the rate of change of prevalence becomes positive. Dots represent individual posterior draws.

## C Sensitivity Analysis

**Figure S10:**
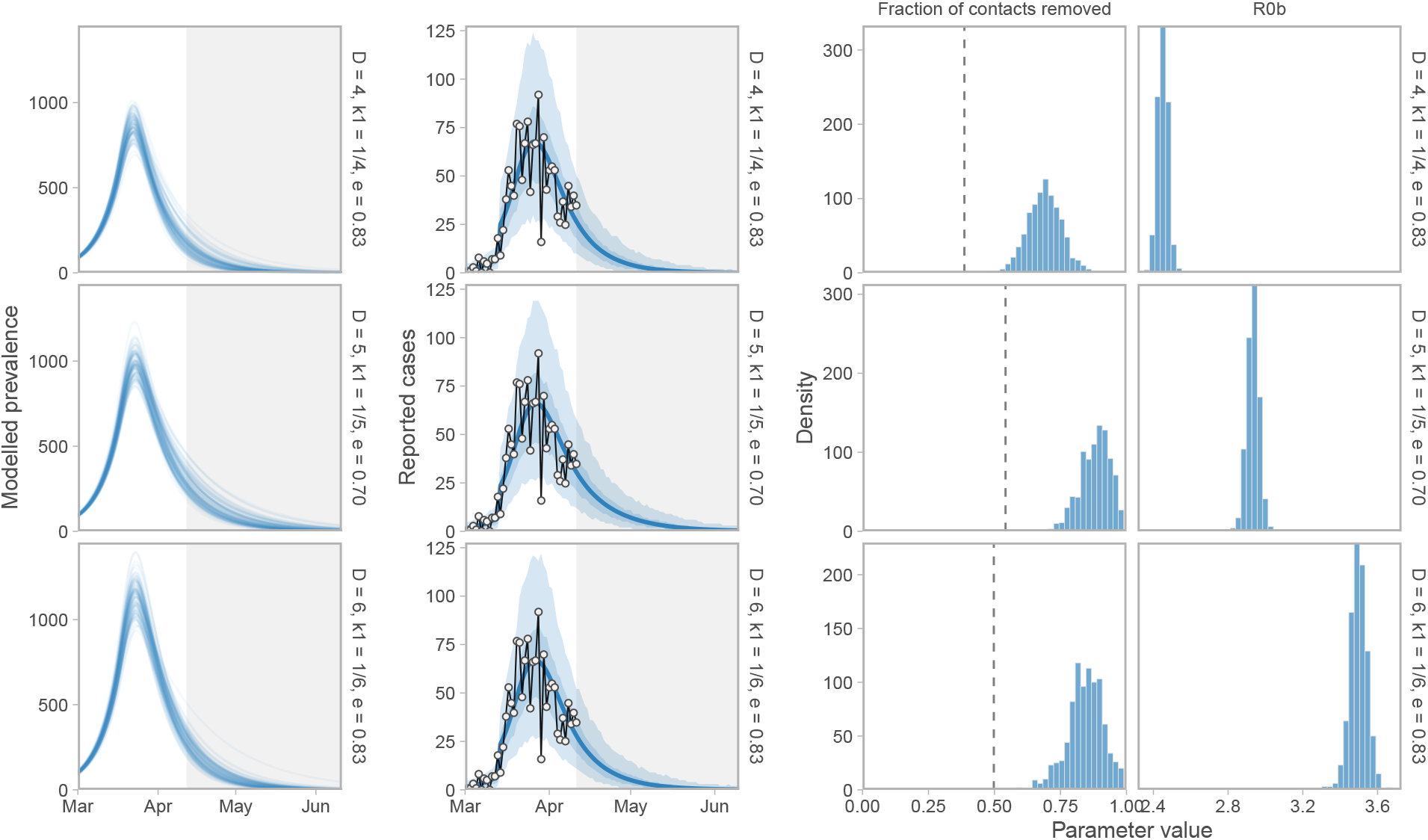
Sensitivity of prevalence and case-count projections, *R*_0b_, and the fraction-of-contacts-removed estimate (1− *f*_2_) to three sets of alternative parameter values: (1) *D* = 4, *k*_1_ = 1 / 4, *u*_*r*_ = 0.2 (shorter duration); (2) *D* = 5, *k*_1_ = 1 / 5, *u*_*r*_ = 0.3/ 0.7 (a lower proportion of people physical distancing: 70% vs. 83% in the main analysis); and (3) *D* = 6, *k*_1_ = 1 / 6, *u*_*r*_ = 0.2 (longer duration). Histograms illustrate the posterior distribution of MCMC samples. Dashed vertical line represents the threshold value below which an exponential increase occurs.

**Figure S11:**
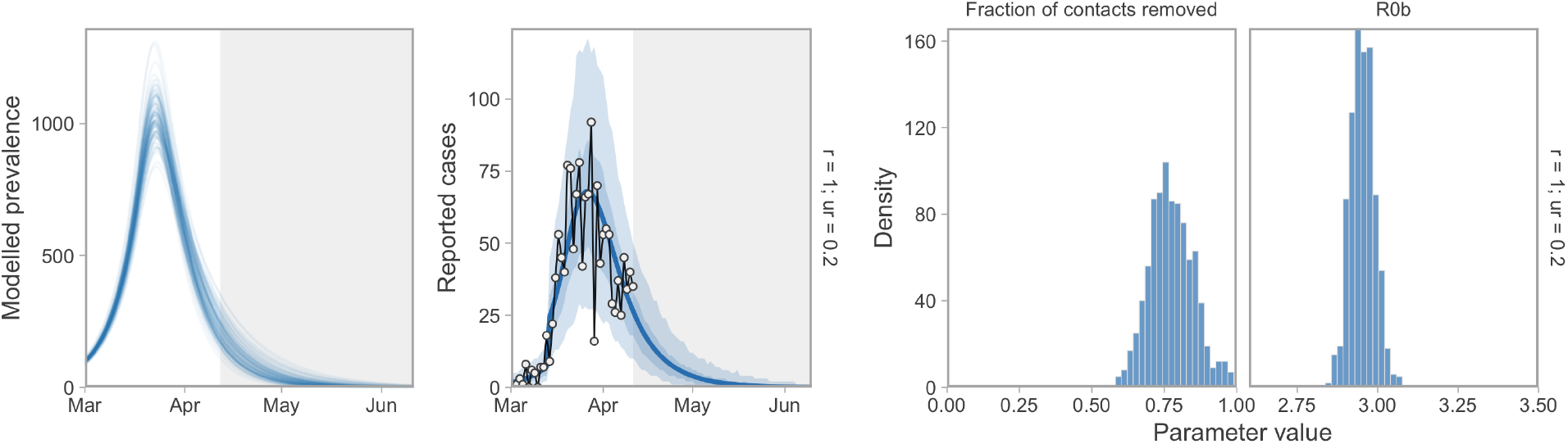
Prevalence and case-count projections, *R*_0b_, and the fraction-of-contacts-removed estimate (1− *f*_2_) with distancing rate parameters *u*_*d*_ = 1 and *u*_*r*_ = 0.2. The ratio *e* is the same as at baseline but these rates are 10 times higher. The model dynamics are sensitive to *e* but not to the choice of rates. Accordingly, these estimates are as in Figure 4.

**Figure S12:**
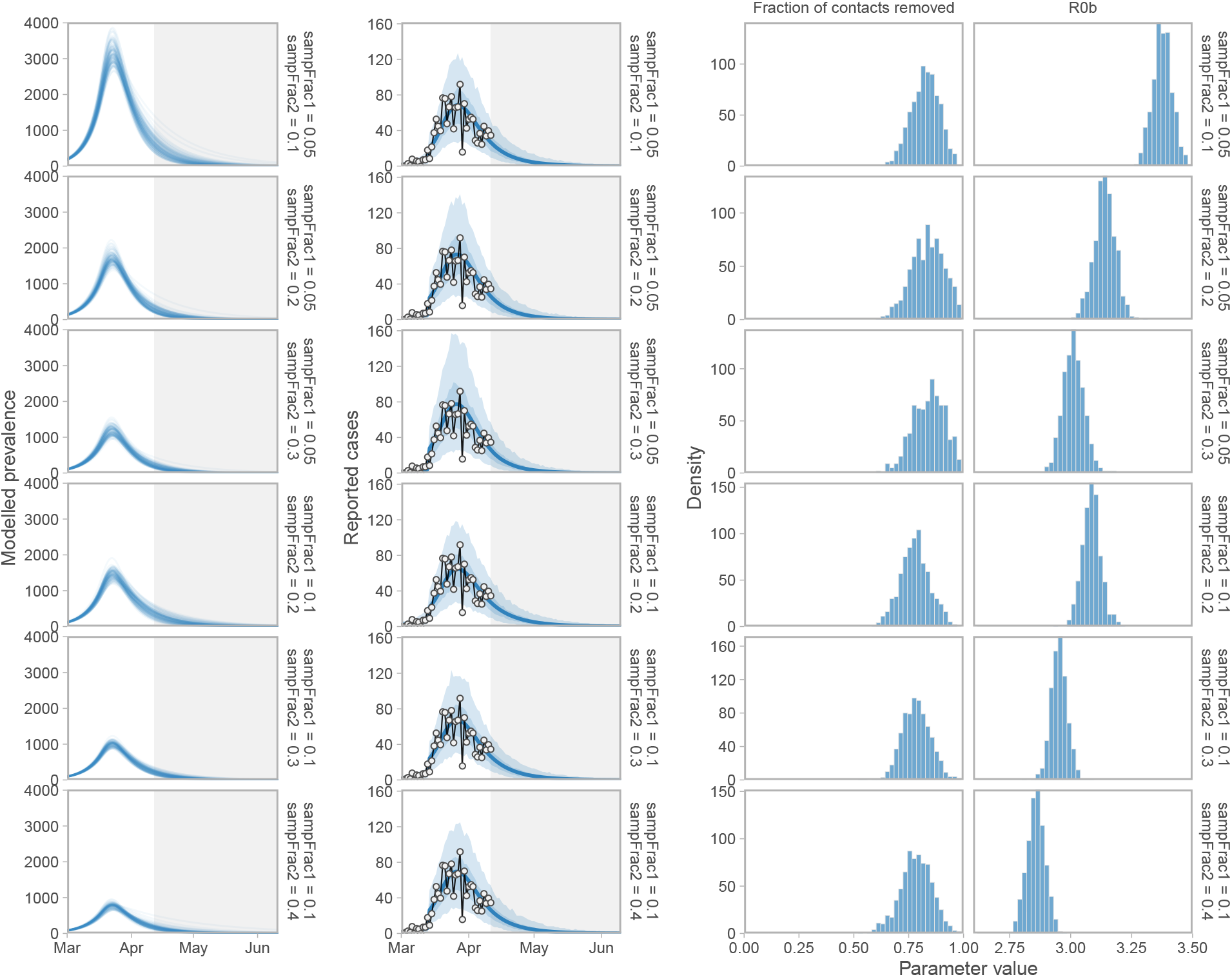
Sensitivity of prevalence and case-count projections, *R*_0b_, and the fraction that contacts are removed (1 − *f*_2_) to assumed fractions of positive cases sampled. “sampFrac1” refers to the assumed sample fraction before March 14, 2020 and “sampFrac2” refers to the assumed sample fraction on and after March 14, 2020. Histograms illustrate the posterior distribution of MCMC samples.

**Figure S13:**
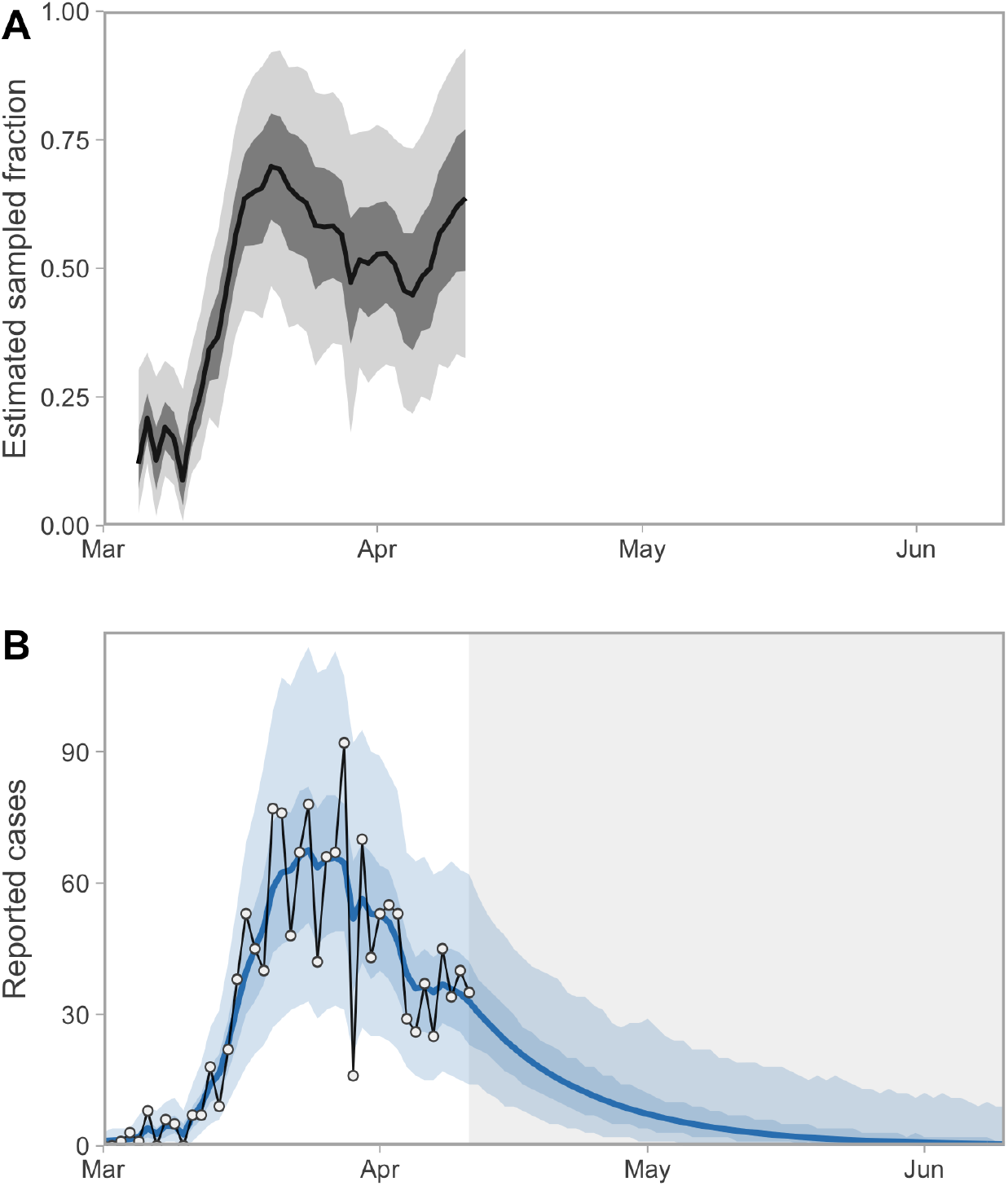
Alternative model fitted with a random walk on the fraction of positive cases sampled. (A) Estimated sampled fraction random walk. The random walk starts on the fifth day (it is fixed at 0.1 before then) and then evolves with a fixed standard deviation of 0.1. We placed a Beta prior on the initial value with a mean and standard deviation of 0.2 and 0.2. (B) Resulting model fit with a sampled-fraction random walk. Shaded ribbons represent 50% and 90% credible intervals. Thick lines represent medians (A) and means (B) of the posterior distribution. Although the shape in panel A is plausible, we deemed the scale of the estimated values to be implausibly high and therefore used a fixed value of 0.3 after day 14 in the main models in this paper.

